# Altered Lateralization of the Cingulum in Deployment-Related Traumatic Brain Injury: An ENIGMA Military-Relevant Brain Injury Study

**DOI:** 10.1101/2022.05.04.22274510

**Authors:** Emily L Dennis, Mary R Newsome, Hannah M Lindsey, Maheen M Adams, Tara A Austin, Seth G Disner, Blessen C Eapen, Carrie Esopenko, Carol E Franz, Elbert Geuze, Courtney Haswell, Sidney R Hinds, Cooper B Hodges, Andrei Irimia, Kimbra Kenney, Inga K Koerte, William S Kremen, Harvey S Levin, Rajendra A Morey, John Ollinger, Jared A Rowland, Randall S Scheibel, Martha E Shenton, Danielle R Sullivan, Leah D Talbert, Sophia I Thomopoulos, Maya Troyanskaya, William C Walker, Xin Wang, Ashley L Ware, J Kent Werner, Wright Williams, Paul M Thompson, David F Tate, Elisabeth A Wilde

## Abstract

Traumatic brain injury (TBI), a significant concern in military populations, is associated with alterations in brain structure and function, cognition, as well as physical and psychological dysfunction. Diffusion magnetic resonance imaging (dMRI) is particularly sensitive to changes in brain structure following TBI, as alterations in white matter (WM) microstructure are common. However, dMRI studies in mild TBI (mTBI) are conflicting, likely due to relatively small samples, sample heterogeneity (demographics, pre- and comorbidities) and injury characteristics (mechanism; chronicity). Furthermore, few studies account for brain asymmetry, which may impact cognitive functions subserved by WM tracts. Examining brain asymmetry in large samples may increase sensitivity to detect heterogeneous areas of subtle WM alteration in mTBI.

Through the Enhancing Neuroimaging and Genetics through Meta-analysis (ENIGMA) Military-Relevant Brain Injury working group, we conducted a mega-analysis of neuroimaging and clinical data from 16 cohorts of Active Duty Service Members and Veterans (*n=*2,598; 2,321 males/277 females; age 19-85 years). 1,080 reported a deployment-related TBI, 480 had a history of only non-military-related TBI, 823 reported no history of TBI, and 215 did not differentiate between military and non-military TBI. dMRI data were processed in a harmonized manner along with harmonized demographic, injury, psychiatric, and cognitive measures. Hemispheric asymmetry of fractional anisotropy (FA, a common proxy for myelin organization) was calculated for 19 WM tracts and compared between those with and without TBI history.

FA in the cingulum showed greater asymmetry in individuals with a history of deployment-related TBI; this effect was driven by greater left lateralization in the group with TBI. There was a trend towards lower FA of the right cingulum in the TBI group. These results remained significant after accounting for potentially confounding variables including posttraumatic stress disorder, depression, and handedness and were driven primarily by individuals who had sustained their worst TBI before age 40. We further found that alterations in the cingulum were associated with slower processing speed and poorer set shifting.

The results indicate an enhancement of the previously reported natural left laterality, possibly due to vulnerability of the non-dominant hemisphere or compensatory mechanisms in the dominant hemisphere. The cingulum is one of the last WM tracts to mature, reaching peak FA around 42 years old. This effect was primarily detected in individuals whose worst injury occurred before age 40, suggesting that the protracted development of the cingulum may lead to increased vulnerability to insults, such as TBI.

## Introduction

Traumatic brain injury (TBI) has been a common battlefield injury throughout history, but has recently received increased attention due to its high prevalence in recent military operations in Iraq and Afghanistan. The Traumatic Brain Injury Center of Excellence reports that Service Members (SM) experienced 450,000 TBIs between 2000-2021,^1^ the majority of which were classified as mild TBIs (mTBIs). While many patients appear to recover from these “invisible injuries”^2^ - typically within weeks - some continue to report somatic, cognitive, and psychological symptoms even several years later.^3^ TBI occurring during deployment has been associated with a greater variety of outcomes than TBI sustained outside deployment.^4–8^ Blast-related TBI in particular has been called the “signature injury” of post-9/11 conflicts (e.g., OEF/OIF/OND). Understanding brain structural changes and their relation to neurobehavioral sequelae following blast-related injuries, however, requires further elucidation as research on mTBI due to impact may not apply as the injury mechanisms differ.

Neuroimaging has revealed persistent alterations in brain structure and function long after other extracranial injuries have healed.^9^ Diffusion magnetic resonance imaging (dMRI) is particularly sensitive to the axonal neuropathology of TBI, but there is mixed evidence for alterations in white matter (WM) organization after injury, especially after mTBI.^10–16^ Heterogeneity among patients may make effects of mTBI especially difficult to detect in group analyses, leading some researchers to take the “pothole” approach, i.e., reporting global WM disruption irrespective of location.^11,12,17^ Larger sample sizes may also partially address the issue of heterogeneity, and big data approaches could facilitate sophisticated modeling to identify patient clusters with different patterns of WM disruption. One prior analysis on a subset of the cohorts included in this analysis used non-negative matrix factorization (NMF), a data-driven approach, to reduce measurements along the WM core (the skeleton) to a set of components that accounted for a large amount of the variance. This analysis revealed an age-dependent effect of TBI on FA (fractional anisotropy, a common proxy for WM integrity), such that the age-related decreases in FA were steeper than expected after TBI.^18^ Here we included the full available sample of size near 2,600.

There are inherent asymmetries in the typically-developed, healthy brain, many of which appear reliably across individuals.^19^ Several WM tracts have well-established asymmetry, such as left lateralization of the cingulum and of the arcuate fasciculus, while asymmetry-related findings for other tracts have been mixed.^20^ This asymmetry may explain some of the functional lateralization of the brain.^21^ We examined asymmetry of lateralized tracts to reduce the number of statistical tests and to be more sensitive to heterogeneity among patients. Asymmetry standardizes measures to the individual, to a degree, making each his/her own reference and thus increasing sensitivity to subtle, heterogeneous effects.^22^

The Enhancing NeuroImaging Genetics through Meta-Analysis (ENIGMA) Military-Relevant Brain Injury working group is part of the broader ENIGMA Brain Injury working group (https://enigma.ini.usc.edu/ongoing/enigma-tbi/), a global collaboration among neuroimaging researchers focusing on various TBI patient populations.^23–25^ It was built on the framework created by the ENIGMA consortium, which seeks to achieve greater statistical power through harmonized image processing and meta/mega-analysis.^26^ The ENIGMA Diffusion Tensor Imaging (DTI) workflow^27^ has been used to identify altered WM organization across a range of disorders, including moderate/severe TBI in pediatric patients,^28^ post-traumatic stress disorder (PTSD),^29^ major depression,^30^ and other conditions.^31^ We used the ENIGMA DTI workflow to analyze asymmetry of WM organization in military brain injury across sixteen cohorts. We hypothesized that there would be some structures displaying greater asymmetry in participants with a history of TBI.

## Materials and Methods

### Study Samples

The ENIGMA Military-Relevant Brain Injury dMRI analysis included 16 cohorts from two countries, totalling 1,775 participants with a history of TBI and 823 comparison participants with no history of TBI. Of the 1,775 participants reporting a history of TBI, 1,080 had at least one deployment-related injury, 480 had only a history of non-military TBI, and 215 did not have information regarding injury context (military vs. non-military). Cohorts included both Veteran and Active Duty SM. While the majority of cohorts focused on OEF/OIF/OND Military SM, two cohorts focused on Vietnam Veterans, thus including a significantly older population. Across the cohorts, the age range was 18-85 years, with an average age of 41.7 ± 12.7 years. **Table 1** provides basic demographic and clinical details on the cohorts, and inclusion and exclusion criteria for each study are in **Supplementary Table 1**. All participants provided written informed consent approved by local institutional review boards. Some cohorts shared raw imaging data with the central site (University of Utah), where they were processed and analyzed, while others processed data locally and shared the summary measures with the central site for mega-analysis.

**Table 1.** Demographic information across cohorts. Numbers are shown for total sample, TBI (any) and control groups, and male/female. The age range, average age in years with standard deviation (SD), sample TBI severity, and the TBI scale collected are also listed.

| Site | Total N | TBI | Control | M | F | Age range | Average age (SD) | Severity | TBI Scale |
| --- | --- | --- | --- | --- | --- | --- | --- | --- | --- |
| **ADNIDoD** | 109 | 67 | 42 | 109 | 0 | 60-85 | 69.6 (4.6) | Mod/Sev | Custom |
| **BETTER/UMCU** | 84 | 12 | 72 | 84 | 0 | 21-57 | 36.2 (9.5) | Mild | Custom |
| **CENC** | 1,028 | 837 | 191 | 891 | 137 | 22-70 | 40.1 (9.8) | Mild | PCE |
| **Chronic Effects** | 80 | 64 | 16 | 72 | 8 | 23-54 | 34.7 (7.4) | Mild | Polytrauma |
| **Duke1** | 105 | 70 | 35 | 86 | 19 | 24-46 | 39.6 (8.8) | Mild | MN-BEST/QCuBE |
| **Duke2** | 84 | 50 | 34 | 58 | 26 | 23-67 | 40.1 (11.3) | Mild | MN-BEST/QCuBE |
| **HDFT** | 28 | 23 | 5 | 23 | 5 | 24-58 | 36.9 (8.7) | Mild-mod | Custom |
| **Houston Blast** | 42 | 32 | 10 | 39 | 3 | 21-54 | 32.4 (8.0) | Mild | Polytrauma |
| **INTRuST** | 85 | 54 | 31 | 69 | 16 | 18-69 | 39.4 (12.8) | Mild | Custom |
| **iSCORE** | 101 | 30 | 71 | 88 | 13 | 19-51 | 35.8 (7.7) | Mild | Custom |
| **NICoE** | 258 | 210 | 48 | 246 | 12 | 19-59 | 39.4 (6.5) | Mild | OSU |
| **Rover Wiser** | 53 | 23 | 30 | 34 | 19 | 21-58 | 31.4 (6.4) | Mild | Polytrauma |
| **SPIRE/GBET** | 53 | 39 | 14 | 47 | 6 | 24-50 | 35.3 (6.9) | Mild-severe | Polytrauma |
| **VA Minneapolis1** | 124 | 92 | 32 | 120 | 4 | 23-62 | 34.3 (8.5) | Mild | MN-BEST |
| **VA Minneapolis2** | 130 | 99 | 31 | 121 | 9 | 22-59 | 32.8 (8.4) | Mild | MN-BEST |
| **VETSA** | 234 | 73 | 161 | 234 | 0 | 56-66 | 61.8 (2.6) | Mild | Custom |
| **Total** | **2,598** | **1,775** | **823** | **2,321** | **277** | **19-85** | **41.7 (12.7)** |  |  |

### Harmonizing Injury Data

Details on injuries were collected using a range of tools across sites (see **Table 1** and **Supplementary Table 2**). From these disparate scales we extracted a number of common variables, not all of which were available for all cohorts: history of TBI (any; Y/N), history of deployment-related TBI (Y/N), history of blast-related TBI (Y/N), number of reported TBIs, history of TBI with loss of consciousness (LOC; Y/N), history of TBI with post-traumatic amnesia (PTA; Y/N), number of TBIs with LOC/PTA, longest reported duration of LOC/PTA, and time since first, worst, and most recent TBI. Longest reported LOC/PTA were largely self-reported and binned into the following categories: LOC - none, <1 min, 1-10 min, 10-30 min, 30 min-1 day, >1 day; PTA - none, <1 hour, 1 hour - 1 day, 1-7 days, >7 days.

### Image Acquisition and Processing

The acquisition parameters for each cohort are provided in **Supplementary Table 3**. Preprocessing, including eddy current correction, echo-planar imaging-induced distortion correction, and tensor fitting, was completed at the University of Utah for those sites sharing raw data, or locally for the others. Recommended protocols and quality control procedures are available on the ENIGMA-DTI and NITRC (Neuroimaging Informatics Tools and Resources Clearinghouse) webpages. These procedures were recommended but harmonization of preprocessing schemes was not enforced across sites to accommodate site- and acquisition-specific pipelines. Once tensors were estimated, they were mapped to the ENIGMA-DTI template, projected onto the ENIGMA-DTI template, and then averaged within regions of interest (ROIs; http://enigma.ini.usc.edu/protocols/dti-protocols/) in a TBSS-based approach.^32^ Further details and ROI abbreviations can be found in **Supplementary Note 1**. In a subset of sites, we extracted motion parameters from the eddy current correction step to determine if motion played a role in our case-control findings. We compared rotation and translation averaged across the X, Y, and Z axes and found no significant differences in motion between groups (all *p*>0.05). We calculated both FA lateralization index and asymmetry for each lateralized ROI:

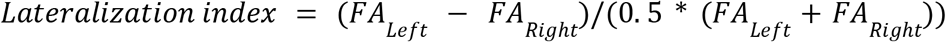

where asymmetry was simply the absolute value of the lateralization index. Significant effects with asymmetry were followed *post hoc* with examinations of the lateralization index. Asymmetry was the primary measure as it would detect alterations irrespective of direction.

### Statistics

As each site shared the individual-level ROI data, mega-analysis was possible. Linear mixed effects models were implemented with *lme* in R 3.1.3. Nested random effects were used to control for cohort and site, as some studies included multiple data collection sites. The average correlation in asymmetry between all ROIs was *r*=0.13. A Bonferroni correction is overly conservative when there are correlations between the multiple dependent measures being tested. Therefore, we followed recent ENIGMA analyses^29^ and calculated the effective number of independent tests based on the observed correlation structure between the alternate responses using two different methods. The equation of Li and Ji^33^ yielded an effective number of tests of V_eff_=16, giving a significance threshold of *p*<0.05/16=0.003125. Results that did not pass correction for multiple comparisons (0.05>*p*>0.003125) are reported for completeness, but not interpreted. Across analyses, Cohen’s *d* statistics are reported for group comparisons and unstandardized betas (*b*) are reported for linear regressions.

### Quadratic Age Term

We first conducted analyses to determine whether a quadratic age term, age^2^, should be included in statistical models along with age and gender, as age has a nonlinear effect on FA.^34^ The effect of this term upon the regression was not significant, so age^2^ was not included in subsequent analyses.

### Primary Group Comparisons

As our focus is military-relevant TBI, our primary analysis compared participants with a history of deployment-related TBI to those with no history of TBI, excluding individuals reporting a history of only non-military TBI and participants whose records did not specify the source of TBI. Deployment-related TBI included both combat and non-combat injuries. As a specificity analysis, we examined group differences between participants with a history of only non-military TBI to non-TBI. We also compared individuals with a history of blast-related TBI to those with no history of TBI, for whom that information was available. *Subgroups*: Two cohorts focused on Vietnam Veterans. These cohorts were significantly older than the other cohorts, generally had more severe injuries, and were far more temporally remote from their injuries. We repeated group analyses without these two cohorts as well. We also conducted group analyses by gender. *Interactions:* We analyzed potential interactions between group and age or gender.

### Injury Variables

We analyzed two other groupings based on injury data: any TBI with LOC and any TBI with PTA. We also examined potential linear relationships with several variables of interest: longest reported LOC/PTA, time since first/worst/most recent injury, and number of TBIs reported.

### Symptom Inventories

Current PTSD and depression symptoms were assessed using different scales across sites yielding categorical variables (Y/N), summarized in **Supplementary Note 2**. For sites that collected versions of the PTSD Checklist (PCL), we were able to calculate a harmonized PCL score based on recent work from our group.^35^ As there was more variety in the depression inventories collected across sites, we did not have a harmonized measure of depression severity. Seven sites collected the Symptom Checklist 90 (SCL-90), Brief Symptom Inventory (BSI), or Neurobehavioral Symptom Inventory (NSI). From these, we extracted two harmonized factors, based on past published work,^36–40^ one for somatic symptoms and one for affective symptoms, detailed in **Supplementary Note 2**.

### Cognitive Function

Across the 16 cohorts included in this study, seven cohorts collected the Trail Making Test (TMT). TMT Part A measures visual search and motor speed, Part B measures set shifting, and subtracting Part A from Part B gives a more specific measure of executive control.^41^ Participants with Part A or Part B completion times greater than 3 SD above the study-wide mean were winsorized to 3 SD above the mean.

### Other Potential Confounds

Three additional potential confounds were also analyzed in our analyses: education, motion, and handedness. We examined whether the number of years of education was associated with asymmetry or was different between the TBI and control groups, and whether motion during the scan was different between TBI and control groups. Details on how motion was assessed may be found in **Supplementary Note 1**. Two sites (Chronic Effects and Rover Wiser) enrolled only right-handed individuals.

### Data Availability

Researchers interested in accessing the data described here must first join the ENIGMA Military-Relevant Brain Injury group and agree to abide by the Memorandum of Understanding governing data sharing and authorship. All projects are opt-in, so the specific cohorts available will differ.

## Results

### Primary Group Comparisons

We found significantly greater FA asymmetry in the group with a history of deployment-related TBI compared to the group with no history of TBI in the cingulum (*d*=0.18, *p*<0.001), along with greater asymmetry in the superior longitudinal fasciculus (SLF), although this did not survive multiple comparisons correction (*d*=0.10, *p*=.027). *Post hoc* analysis of the cingulum revealed greater left lateralization in the TBI group (*d*=0.15, *p*=.001; **Figure 1**). We further examined group differences in the FA of the left and right cingula and found a trend toward lower FA of the right cingulum in the TBI group (*d=*-0.07, *p*=.11). The higher asymmetry was also present in the blast-related TBI vs. no TBI comparison, although it did not survive correction for multiple comparisons (*d*=0.11, *p*=.019). CENC was the only cohort with a variable for (ostensibly) pure blast-related TBI, so as an exploratory analysis we examined cingulum asymmetry between individuals with history of pure blast-related TBI compared to individuals with no TBI history, finding no significant group difference in cingulum asymmetry (*d*=-0.06, *p*=.226). In a *post hoc* analysis of the cingulum, the differences in the deployment-related TBI group remained when covarying for current PTSD (*d*=0.16, *p*<0.001), current depression (*d*=0.16, *p*=.001), and both current PTSD and current depression (*d*=0.15, *p*=0.002). Comparing individuals with a history of only non-military TBI to the non-TBI group, the cingulum effect was not significant (*d*=0.08, *p*=.134), suggesting that our result is specific to deployment-related TBI and not mTBI in general. These results are summarized in **Table 2** and **Supplementary Table 4**.

**Table 2.** Group differences in tract asymmetry. Results from group analyses are shown comparing deployment-related TBI to no TBI, blast-related TBI to no TBI, and pure blast TBI to no TBI, along with total sample size for each comparison. Cohen’s *d*, 95% CI, and uncorrected *p*-values are shown, **bolded** statistics are those that survive correction for multiple comparisons, *italicized* statistics are those that do not survive multiple comparisons correction (0.05>*p*>0.003125). Region of interest (ROI) abbreviations: anterior corona radiata (ACR), anterior limb of internal capsule (ALIC), cingulum (CGC), hippocampal cingulum (CGH), corticospinal tract (CST), corona radiata (CR), external/extreme capsule (EC), fornix-stria terminalis (FX/ST), internal capsule (IC), posterior corona radiata (PCR), posterior limb of internal capsule (PLIC), posterior thalamic radiation (PTR), retrolenticular limb of internal capsule (RLIC), superior corona radiata (SCR), superior fronto-occipital fasciculus (SFO), superior longitudinal fasciculus (SLF), sagittal stratum (SS), tapetum (TAP), uncinate (UNC).

| ROIs | Deployment-related TBI<br>(*n*=1,872) |  | Blast-related TBI<br>(*n*=1,509) |  | Pure blast TBI (*n*=337) |  | HX of TBI with LOC |  |
| --- | --- | --- | --- | --- | --- | --- | --- | --- |
|  | *d* [CI] | *p* | *d* [CI] | *p* | *d* [CI] | *p* | *d* [CI] | *p* |
| **ACR** | -0.01 [-0.11, 0.08] | 0.75 | -0.02 [-0.11, 0.07] | 0.63 | -0.06 [-0.16, 0.03] | 0.17 | 0.06 [-0.04, 0.15] | 0.24 |
| **ALIC** | 0.04 [-0.05, 0.13] | 0.35 | 0.03 [-0.06, 0.12] | 0.52 | 0.05 [-0.04, 0.14] | 0.32 | 0.05 [-0.04, 0.14] | 0.29 |
| **CGC** | **0.18 [ 0.08, 0.27]** | **0.00015** | *0.11 [ 0.02, 0.20]* | *0.019* | -0.06 [-0.15, 0.04] | 0.23 | 0.06 [-0.04, 0.15] | 0.25 |
| **CGH** | 0.01 [-0.08, 0.10] | 0.81 | 0.01 [-0.08, 0.10] | 0.82 | -0.04 [-0.13, 0.05] | 0.39 | 0.02 [-0.07, 0.11] | 0.66 |
| **CST** | -0.08 [-0.17, 0.01] | 0.086 | -0.05 [-0.14, 0.04] | 0.24 | -0.07 [-0.16, 0.02] | 0.12 | -4.1e-3 [-0.09, 0.09] | 0.93 |
| **CR** | 0.06 [-0.03, 0.15] | 0.2 | 0.04 [-0.05, 0.13] | *0.41* | 0.04 [-0.05, 0.14] | 0.34 | *0.12 [ 0.03, 0.21]* | *0.009* |
| **EC** | -0.04 [-0.13, 0.05] | 0.42 | -0.02 [-0.11, 0.07] | 0.73 | 0.09 [ 0.00, 0.18] | 0.056 | -0.04 [-0.13, 0.05] | 0.39 |
| **FX/ST** | 0.03 [-0.06, 0.12] | 0.46 | 0.02 [-0.07, 0.11] | 0.62 | -0.07 [-0.16, 0.02] | 0.14 | 0.03 [-0.06, 0.12] | 0.54 |
| **IC** | 0.05 [-0.04, 0.14] | 0.3 | 0.04 [-0.05, 0.13] | 0.39 | *0.11 [ 0.02, 0.20]* | *0.017* | 0.02 [-0.07, 0.11] | 0.68 |
| **PCR** | 0.04 [-0.05, 0.13] | 0.39 | 0.04 [-0.05, 0.13] | 0.42 | 0.02 [-0.07, 0.12] | 0.6 | 0.06 [-0.03, 0.16] | 0.17 |
| **PLIC** | 1.0e-3 [-0.09, 0.09] | 0.98 | -0.01 [-0.10, 0.08] | 0.78 | 5.1e-3 [-0.09, 0.10] | 0.91 | -0.03 [-0.12, 0.06] | 0.46 |
| **PTR** | 0.07 [-0.02, 0.16] | 0.12 | 0.06 [-0.03, 0.15] | 0.19 | 0.05 [-0.04, 0.14] | 0.27 | 0.07 [-0.02, 0.16] | 0.15 |
| **RLIC** | 3.0e-3 [-0.09, 0.09] | 0.95 | 0.02 [-0.07, 0.11] | 0.71 | 0.08 [-0.01, 0.17] | 0.088 | 0.04 [-0.05, 0.13] | 0.42 |
| **SCR** | 0.06 [-0.03, 0.15] | 0.22 | 0.09 [ 0.00, 0.18] | 0.055 | 0.05 [-0.04, 0.14] | 0.31 | -1.5e-3 [-0.09, 0.09] | 0.97 |
| **SFO** | -0.03 [-0.12, 0.06] | 0.55 | 0.02 [-0.07, 0.11] | 0.69 | 0.02 [-0.07, 0.11] | 0.73 | 0.04 [-0.06, 0.13] | 0.44 |
| **SLF** | *0.10 [ 0.01, 0.19]* | *0.027* | 0.06 [-0.03, 0.15] | 0.18 | 0.03 [-0.06, 0.12] | 0.57 | **0.13 [ 0.05, 0.23]** | **0.0018** |
| **SS** | 0.07 [-0.02, 0.16] | 0.13 | 0.05 [-0.04, 0.14] | 0.26 | -0.06 [-0.15, 0.04] | 0.24 | 0.06 [-0.03, 0.15] | 0.18 |
| **TAP** | -0.08 [-0.18, 0.01] | 0.068 | -0.02 [-0.11, 0.07] | 0.63 | 0.03 [-0.06, 0.12] | 0.53 | -0.02 [-0.11, 0.07] | 0.66 |
| **UNC** | 0.02 [-0.07, 0.11] | 0.67 | 0.02 [-0.07, 0.11] | 0.67 | 0.03 [-0.06, 0.13] | 0.45 | 0.04 [-0.05, 0.13] | 0.35 |

**Figure 1.**
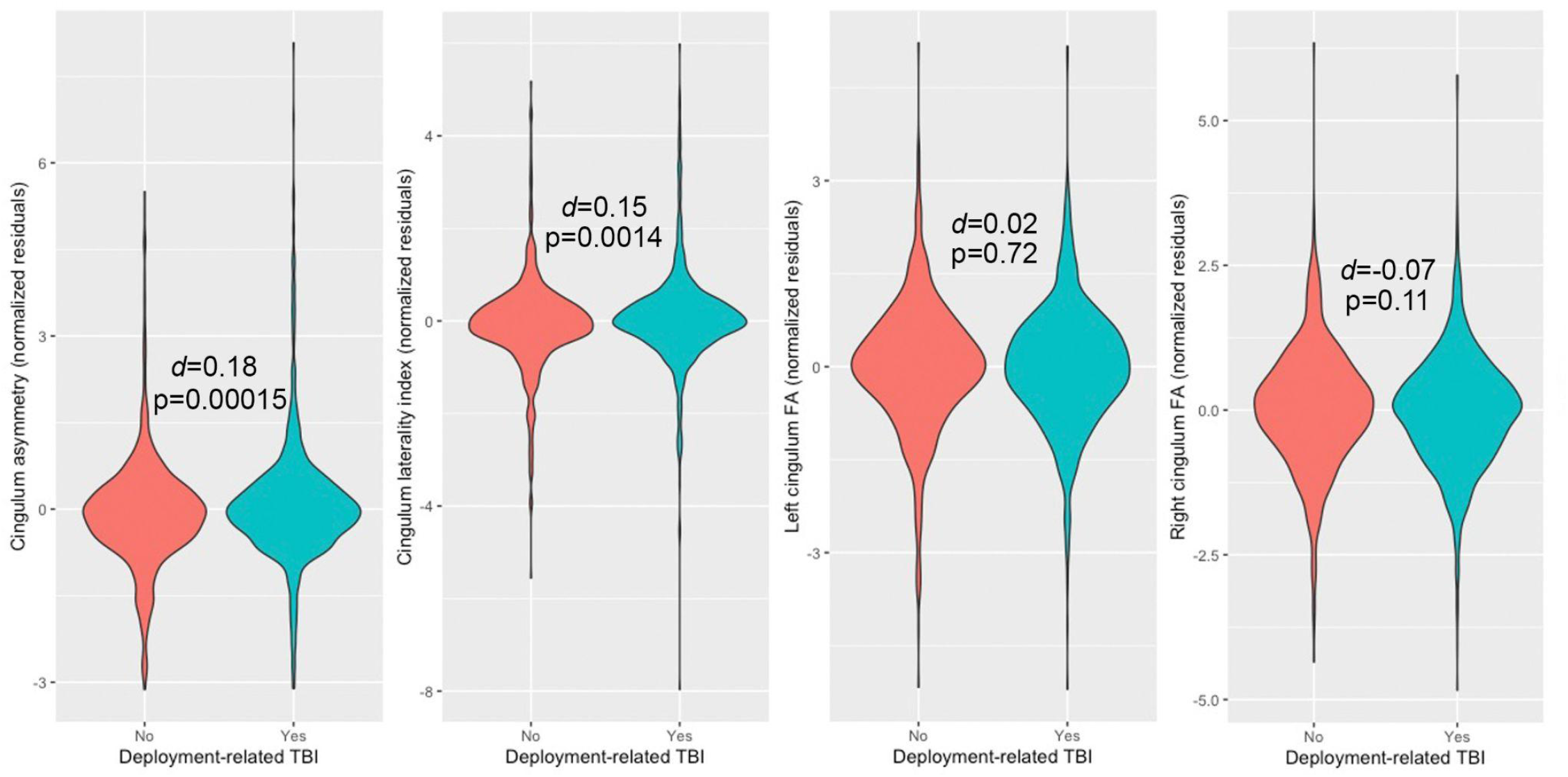
Group differences in cingulum asymmetry. Violin plots are shown for the no-TBI group (red), and deployment-related TBI group (blue) with group *t*-test *p*-values. The values on the *y*-axis are the normalized residuals for cingulum asymmetry, laterality index, left FA, and right FA, accounting for age, gender, and the nested random effects of cohort and site. As the cingulum is generally left-lateralized, actual laterality index values are generally positive, but that is not reflected here due to the adjustments and normalization.

### Subgroups

When the two Vietnam Veteran cohorts were excluded, cingulum results remained significant (*d*=0.17; *p*<.001). Among Vietnam Veterans, differences in the cingulum were not significant (*d*=0.25, *p*=.051), but there was significantly greater asymmetry in the SLF of participants with a history of deployment-related TBI (*d*=0.39, *p*=.003), although this was in a much smaller sample (*n*=253). The cingulum is one of the last tracts to mature^42^, which suggests that it exhibits prolonged vulnerability to environmental insults such as injury. To test this hypothesis, we examined group differences *post hoc* in (A) individuals whose worst injury occurred when they were younger than 40 separately from (B) those whose worst injury occurred when they were older than 45, excluding the two samples with Vietnam Veterans. The cingulum effect remained significant among those injured at a younger age (*d*=0.12, *p*=.017). It was not significant (while the SLF effect was) among those injured at an older age (*d*=0.01, *p*=.83). When males and females were separated, the cingulum effect remained significant in males (*d*=0.17, *p*<.001), but not in females (*d*=0.26, *p*=.077). These results are summarized in **Supplementary Table 4**.

### Interactions

There were no significant interactions between diagnosis (based on deployment-related TBI history) and age or gender. These results are summarized in **Supplementary Table 5**.

### Injury variables

***LOC/PTA:*** *Loss of consciousness (LOC)*: There was significantly more asymmetry in the SLF in participants who had experienced at least one TBI with LOC (*d*=0.13, *p*=.0018, **Figure 2**), as well as in the corticospinal tract (CST), although this association did not survive correction for multiple comparisons (*d*=0.11, *p*=0.0090). *Post hoc* analyses revealed greater right lateralization in the SLF of participants in the LOC group. Examining the FAs of the left and right SLF did not reveal significant results. Longer LOC (longest reported) was associated with greater asymmetry in the uncinate fasciculus (UF), although this did not survive correction for multiple comparisons (*n*=646; *b*=0.0077, *p*=.046). A larger number of reported TBIs with LOC was associated with greater asymmetry in the CST, although this did not survive correction for multiple comparisons (*n*=2,058; *b*=0.0025, *p*=.034). *Post-traumatic amnesia (PTA)*: There were no significant associations between participants who had experienced at least one TBI with PTA and participants with no TBI history. Longer duration of PTA (longest reported) was associated with greater asymmetry in the CGH (hippocampal cingulum), superior *corona radiata* (SCR), SLF, and with lower asymmetry in the PCR, although these did not survive correction for multiple comparisons (*n=*1,028; *b*=0.0087, *p*=.022; *b*=0.0043, *p*=.009; *b*=0.0053, *p*=.008; *b*=-0.0044, *p*=0.036, respectively). Greater number of TBIs with PTA was associated with greater asymmetry in the CST, although this did not survive correction for multiple comparisons (*n*=1,577; *b*=0.0036, *p*=0.0060). ***Time since injury***: None of the associations with time since injury survived multiple comparisons corrections. *Time since first injury*: There was a negative association with asymmetry of the UF (*n*=1,519; *b*=-2.8×10^−4^, *p*=0.047), and positive associations with asymmetry of the posterior *corona radiata* (PCR) and SLF (*b*=1.0×10^−4^, *p*=0.049; *b*=1.2×10^−4^, *p*=0.016, respectively). *Time since worst injury*: There was a negative association with asymmetry of the superior fronto-occipital fasciculus (SFOF; *n*=2,266; *b*=-2.9×10^−4^, *p*=0.021) and a positive association with asymmetry of the SLF (*b*=1.4×10^−4^, *p*=0.025). *Time since most recent injury*: There was a negative association with asymmetry of the internal capsule (IC) and SFOF (*n*=2,238, *b*=-2.0×10^−4^, *p*=.041; *b*=-2.9×10^−4^, *p*=.033, respectively), and a positive association with the asymmetry of the PCR and SLF (*b*=1.2×10^−4^, *p*=.034; *b*=1.4×10^−4^, *p*=.018, respectively). ***Number of TBIs***: There were no significant relationships with number of TBIs (general) or number of blast-related TBIs. These results are summarized in **Supplementary Table 5**.

**Figure 2.**
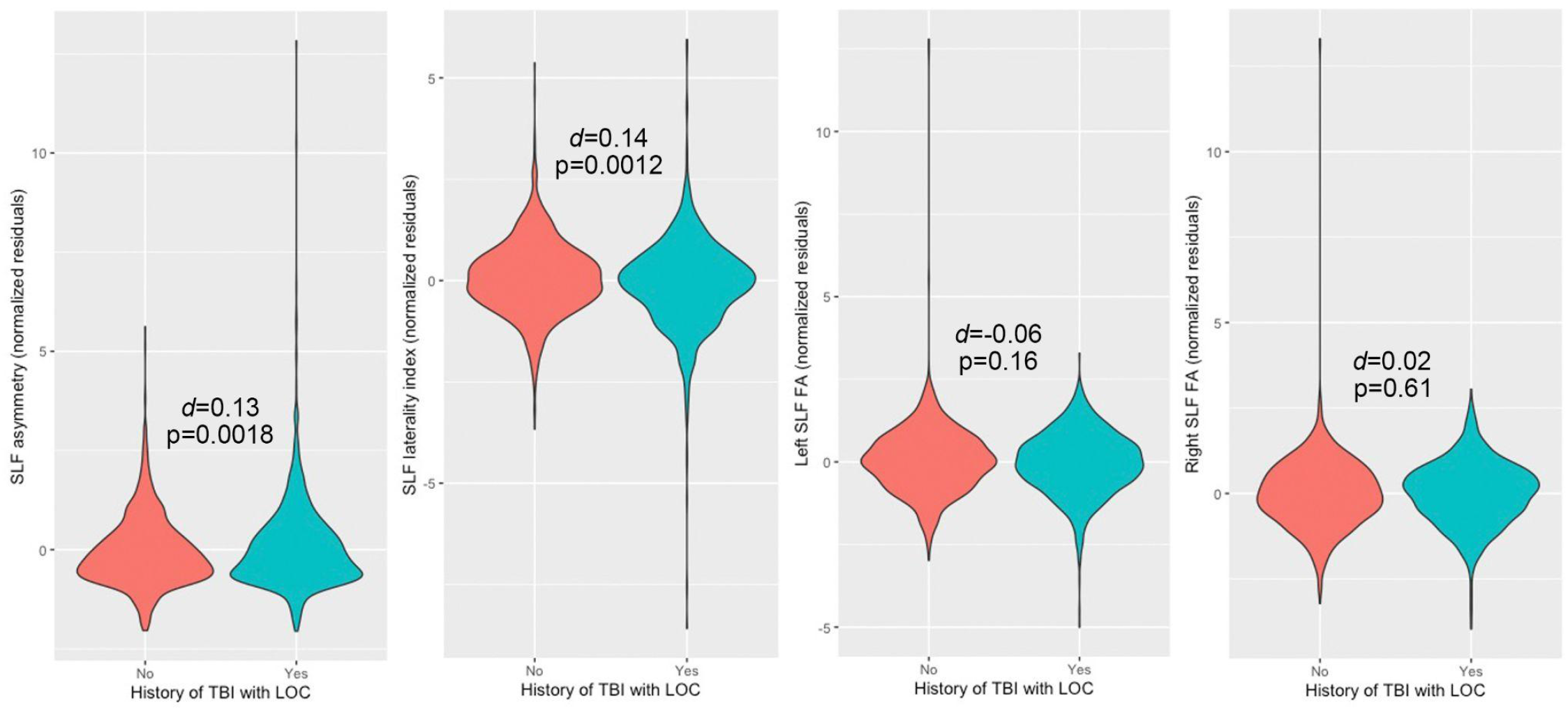
Group differences (based on LOC) in SLF asymmetry. Violin plots are shown for the no-TBI group (red), and TBI with loss of consciousness (LOC) group (blue) with group *t*-test *p*-values. The values on the *y*-axis are the normalized residuals for SLF asymmetry, laterality index, left FA, and right FA (left to right panels), accounting for age, gender, and the nested random effects of cohort and site.

### Cognitive Function

Seven of 16 cohorts in our sample collected the Trail Making Test (TMT, parts A and B), including 1,613 participants, 676 of whom had a history of deployment-related TBI. In the no-TBI group (*n*=506), there were no detectable significant associations between cingulum FA and TMT performance. We examined the cingulum effect *post hoc* to test whether there were functional consequences of the shift in asymmetry. There were no significant associations with asymmetry in the deployment-related TBI group. There were borderline negative associations in the deployment-related TBI group between the lateralization index of the cingulum and TMT-A and TMT-B time (*b*=-5.5×10^−4^, *p*=.071; *b*=-1.8×10^−4^, *p*=.082, respectively, **Supplementary Figure 1**). Within the deployment-related TBI group, the FA of the left cingulum was negatively associated with TMT-A, TMT-B, and TMT-BsubA (*b*=-5.0×10^−4^, *p*=.008; *b*=-1.7×10^−4^, *p*=.011; *b*=-2.0×10^−4^, *p*=.016, respectively, **Figure 3**). When covarying for handedness, only the association with TMT-A remained (*b*=-5.9×10^−4^, *p*=.032), but the sample was also much smaller (*n*=331).

**Figure 3.**
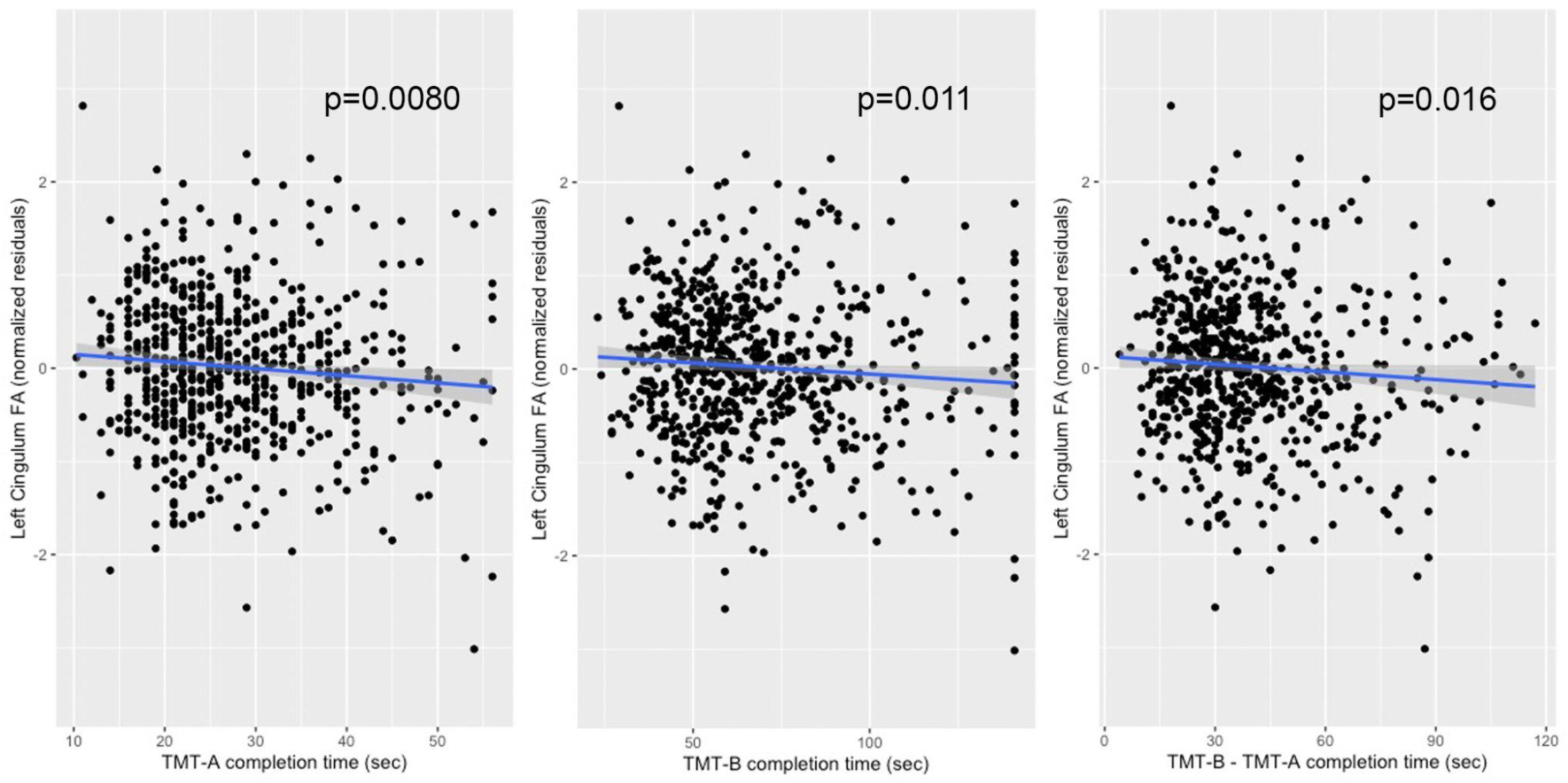
Associations between cingulum microstructure and Trail Making Task (TMT) performance. The three panels (left to right) show significant associations between TMT-A, TMT-B, and TMT-BsubA completion time (in seconds) and left cingulum FA (normalized residuals accounting for age, gender, and the nested random effects of cohort and site). *P*-values are shown for all three regressions. Linear trend line shown with 95% confidence interval, calculated in R 3.6.0. Outliers (>3SD) were removed from the plots (*n*=2).

### Symptoms

*Somatic symptoms*: Examining somatic symptoms (*n*=1,446), we found significantly greater asymmetry in the UF with greater symptom burden (*b*=0.055, *p*<.001). *Post hoc* analyses did not reveal specific lateralization (**Figure 4**). We also found greater asymmetry in the anterior *corona radiata* (ACR), but this did not survive correction for multiple comparisons (*b*=0.012, *p*=.024). *Affective symptoms*: Examining affective symptoms (*n*=1,447), we found greater asymmetry in the ACR, SS, and UF with greater symptom burden, but these did not survive correction for multiple comparisons (*b*=0.0091, *p*=.018; *b*=0.029, *p*=.009; *b*=0.010, *p*=0.041, respectively). *PTSD*: We examined current PTSD both as a categorical variable (Y/N) and a continuous variable, for those sites that collected the PCL. For sites that did, a cutoff score of 33 was used to create a categorical variable. Current PTSD was associated with lower asymmetry in the SCR and higher asymmetry in the sagittal stratum (*n*=2,546; *d*=-0.08, *p*=.035; *d*=0.11, *p*=.004, respectively) although neither of these survived corrections for multiple comparisons. Eight of 16 cohorts collected a version of the PCL, which we harmonized as detailed elsewhere.^35^ Current PTSD severity (within the past month) was associated with higher asymmetry in the UF and sagittal stratum (*n*=1,791; *b*=3.2×10^−4^, *p*=.0036; *b*=1.8×10^−4^, *p*=0.0034, respectively), although neither of these survived corrections for multiple comparisons. *Depression*: We examined current depression as a categorical variable (Y/N), finding no significant associations with asymmetry. These results are summarized in **Table 3**.

**Table 3.** Associations between tract asymmetry and symptoms. Linear regressions are shown for associations with somatic, affective, and posttraumatic stress disorder (PTSD) symptoms, with unstandardized βs and uncorrected *p*-values. Cohen’s *d* with 95% CI are shown for PTSD and depression categorical variables. Sample sizes are shown for all analysis. **Bolded** *p*-values are those that survive correction for multiple comparisons, *italicized p*-values are those that do not survive multiple comparisons correction (0.05>*p*>0.003125). Region of interest (ROI) abbreviations can be found in the legend for **Table 2**.

| ROIs | Somatic symptoms<br>($n=1,446$) | Affective symptoms<br>($n=1,447$) | PTSD (Yes/No,<br>$n=2,546$) | PTSD severity<br>($n=1,791$) | Depression<br>(Yes/No, $n=2,208$) |
| --- | --- | --- | --- | --- | --- |
| ACR | 0.012 (0.024) | 0.0091 (0.018) | 1.3e-3 [-0.08, 0.08] | 8.6e-5 (0.22) | 0.02 [-0.07, 0.10] |
| ALIC | -1.4e-4 (0.98) | 0.0026 (0.53) | 0.03 [-0.05, 0.11] | 3.4e-5 (0.48) | -5.1e-3 [-0.09, 0.08] |
| CGC | 0.0095 (0.31) | -2.9e-4 (0.97) | 0.04 [-0.04, 0.12] | 1.4e-5 (0.87) | 0.03 [-0.05, 0.11] |
| CGH | 0.0090 (0.33) | 0.0087 (0.18) | 0.02 [-0.06, 0.10] | 9.2e-5 (0.31) | 0.03 [-0.05, 0.11] |
| CST | -0.0033 (0.30) | -0.0015 (0.50) | -0.05 [-0.13, 0.03] | -1.7e-5 (0.48) | -0.03 [-0.11, 0.05] |
| CR | 0.0062 (0.50) | 0.0030 (0.66) | -0.01 [-0.09, 0.07] | 2.2e-5 (0.81) | -2.6e-3 [-0.09, 0.08] |
| EC | -0.0046 (0.40) | -0.0015 (0.69) | -7.1e-3 [-0.08, 0.07] | -6.7e-5 (0.11) | -0.02 [-0.11, 0.06] |
| FX/ST | 0.011 (0.14) | 0.0058 (0.28) | 0.07 [-0.01, 0.15] | 3.8e-5 (0.52) | 0.05 [-0.03, 0.13] |
| IC | -8.3e-4 (0.82) | -7.8e-4 (0.76) | -4.0e-3 [-0.08, 0.07] | 4.9e-6 (0.86) | 0.03 [-0.05, 0.11] |
| PCR | -9.1e-4 (0.86) | -0.0018 (0.63) | 0.04 [-0.04, 0.12] | -1.6e-5 (0.68) | 7.7e-3 [-0.08, 0.09] |
| PLIC | 0.0035 (0.57) | -6.9e-4 (0.87) | -6.3e-4 [-0.08, 0.08] | 2.9e-5 (0.52) | 0.02 [-0.06, 0.11] |
| PTR | 0.0069 (0.26) | 0.0024 (0.58) | 0.05 [-0.03, 0.13] | 4.4e-5 (0.36) | 1.1e-4 [-0.08, 0.08] |
| RLIC | -0.0027 (0.61) | -0.0034 (0.37) | -2.3e-3 [-0.08, 0.08] | 1.8e-6 (0.96) | -0.04 [-0.13, 0.04] |
| SCR | -0.0046 (0.27) | -0.0050 (0.092) | *-0.08 [-0.16, -0.01]* | -3.8e-5 (0.22) | -0.03 [-0.11, 0.05] |
| SFO | -0.0042 (0.62) | -0.0039 (0.52) | -0.02 [-0.10, 0.06] | -1.0e-4 (0.29) | 4.0e-3 [-0.08, 0.09] |
| SLF | 8.8e-4 (0.86) | -9.2e-4 (0.79) | -0.03 [-0.11, 0.05] | -2.2e-5 (0.52) | -0.01 [-0.09, 0.07] |
| SS | 0.013 (0.076) | 0.011 (0.041) | *0.11 [0.04, 0.19]* | *1.8e-4 (0.0034)* | 0.07 [-0.01, 0.16] |
| TAP | 0.0051 (0.62) | 0.0046 (0.53) | -5.2e-3 [-0.08, 0.07] | 8.0e-7 (0.99) | -0.06 [-0.14, 0.02] |
| UNC | **0.055 (0.00040)** | 0.029 (0.009) | 0.04 [-0.03, 0.12] | *3.2e-4 (0.0036)* | 0.03 [-0.05, 0.12] |

**Figure 4.**
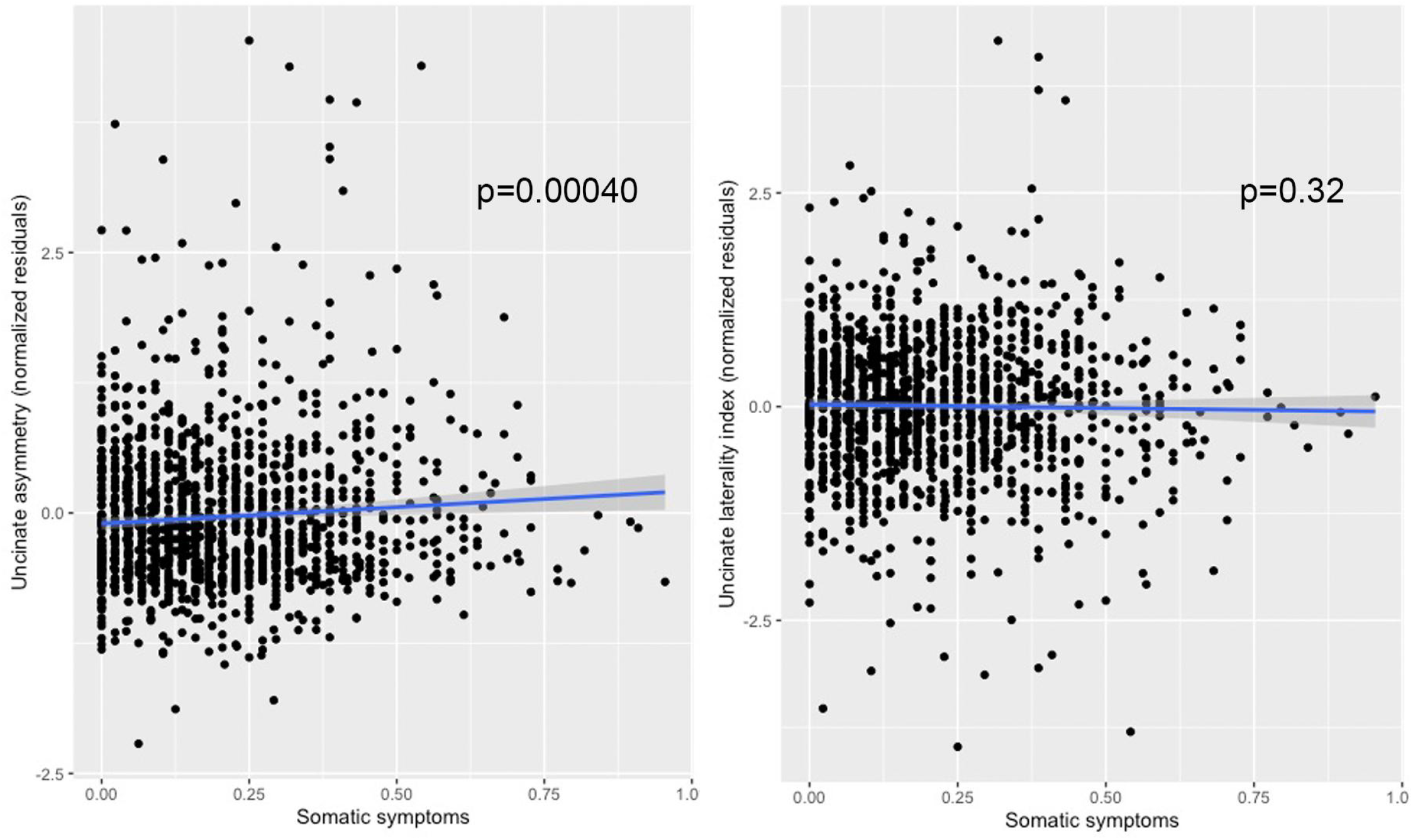
Associations between uncinate asymmetry and somatic symptoms. The left panel shows the significant association between somatic symptoms and uncinate asymmetry (normalized residuals accounting for age, gender, and the nested random effects of cohort and site). The right panel shows the null association between somatic symptoms and uncinate laterality index (normalized residuals accounting for age, sex, and the nested random effects of cohort and site). *P*-values are displayed for both regressions. Linear trend line shown with 95% confidence interval, calculated in R 3.6.0. Extreme outliers (>5SD) were removed from the plots (*n*=5).

### Potential Confounds

#### Education

Years of education did not differ between the TBI and control groups and was not associated with tract asymmetry. *Motion*: Motion parameters were not significantly in individuals with a history of deployment-related TBI. As neither of these variables was significantly related to our measures of interest, they were not included in any analyses. *Handedness*: Handedness was not significantly associated with cingulum asymmetry (*n*=1,876, *d*=0.02, *p*=.73). When covarying for handedness, the group difference in the cingulum remained significant (*n*=1,351, *d*=0.20, *p*<.001). When limiting the analysis to only right-handed individuals, the group difference remained significant (618 v. 565, *d*=0.23, *p*<.001; see **Supplementary Table 4)**.

## Discussion

We examined WM asymmetry in 2,598 military SM and Veterans, reporting a primary finding of greater asymmetry in the cingulum among individuals with a history of deployment-related TBI compared to those with no lifetime history of TBI. Greater asymmetry in the cingulum was driven by greater left lateralization in the group with TBI. The cingulum is a key structure in the limbic system, connecting medial aspects of the frontal and parietal lobes (not including the hippocampal cingulum, as the dorsal and hippocampal were distinct in this study)^43^ and supporting a number of important executive functions, including working memory, inhibition, and processing speed.^44^ The cingulum is also one of the last tracts to mature, with an average age-at-peak FA of 42 years old.^42^ This prolonged maturation may render the cingulum more vulnerable to environmental impacts, and prior structural equation modeling did not show significant genetic contributions to cingulum asymmetry, supporting the hypothesis that environment plays an outsized role.^45^ This age-at-peak happens to coincide with the average age of our sample (41.7 years). Indeed, when we ran the analysis after separating participants whose worst injury occurred before age 40 versus those whose worst injury occurred after age 45 only the comparison in the younger group remained significant. This analysis excluded Vietnam Veterans as the remoteness of their injuries may mean different injury- and age-related processes are occurring. Heterogeneity across cohorts, and the general reliance on self-reports of injuries, including their timing and severity, suggest that this result must be interpreted with caution, but this does present a hypothesis that can be addressed more directly in individual studies. As the sample size differed greatly between these analyses (855 younger TBI participants as opposed to 106 older TBI participants), we cannot rule out the decrease in statistical power as the reason for the difference; however, this result does lend support to the interpretation that the cingulum is particularly vulnerable to injury because of its long maturation.

The length, orientation, and proximity of the cingulum to the falx and the ventricles may contribute to strain and shear forces on the tract during impact.^46–48^ Disentangling the effects of blast-related versus impact-related TBI is difficult, especially in our analysis of heterogeneous, legacy datasets. The differences between blast-related TBI and no TBI were not significant after multiple comparisons correction and the effect size was smaller (*d*=0.18 vs. *d*=0.11), but the sample size was smaller (1,080 vs. 734 TBI), and blast-related injuries in military settings often also involve a co-occurring impact. The CENC cohort was the only one to collect information on TBIs that were due primarily to blast. An exploratory analysis in this small subset of the data (146 vs. 191) did not yield significant results for the cingulum, lending support to the interpretation that the acceleration/deceleration/rotational forces of TBI may affect the cingulum more than the pressure forces of blast-related TBI. Targeted analysis in animal studies and well-characterized cohort studies are necessary to confirm this finding.

Left lateralization of the FA of the cingulum is a well-documented phenomenon in healthy (primarily right-handed) individuals, ^20,49,50^ and is linked with cingulum cognitive functions.^51^ In tracts that are more asymmetric, the non-dominant hemisphere may be more susceptible to damage, as neural resources may be primarily diverted to maintaining integrity of the dominant hemisphere. Lower FA in the right cingulum may be due to less myelination, or to less organized or less densely packed fibers. A large study of young adults using multi-shell dMRI recently showed that the neuronal density of the left cingulum was greater than that of the right cingulum, while the orientation dispersion index (ODI; higher values indicate less coherence in fiber direction) was higher in the right cingulum.^52^ Lower fiber density could mean greater elasticity and thus more stretch during impact; finite element modeling consistently indicates high axonal strain in the cingulum.^53–55^ Some studies have found lower tract asymmetry in women than in men,^45,56^ although such differences may be minor.^20,49^ Less asymmetry may partially explain the lack of significant group differences in females, but the much smaller female sample size (*n*=199) means poorer statistical power and thus limits our ability to draw any conclusions from the female-only analyses.

Alterations in cingulum microstructure have been reported previously in mild TBI,^57–61^ including military-specific mTBI,^10,62,63^ as well as across psychiatric disorders.^31,64,65^ In studies with TBI patients, several have reported associations between WM microstructural organization in the right cingulum and cognitive function, including reaction time and set shifting.^58,66,67^ Prior studies have shown that deployment-related TBI is associated with poorer TMT performance, even when compared to non-deployment TBI.^8^ Altered asymmetry of the cingulum in our sample appeared to be primarily driven by deficits in the right cingulum, leading to an exaggerated laterality index. Lower FA of the left cingulum was linked with slower processing speed and set shifting, as measured by the TMT in the TBI group. This association was not present in the comparison group reporting no history of TBI. Other studies have reported that alterations in WM asymmetry are associated with cognitive deficits.^68^ Two prior studies examined WM asymmetry in mTBI and reported increased asymmetry in the corticospinal tract.^69,70^ Unfortunately this midline tract was not separated into left and right components in the ENIGMA atlas, so we did not have the opportunity to replicate that finding.

We found alterations in the asymmetry of the SLF in individuals who had experienced at least one TBI with LOC. History of TBI with LOC was associated with greater right lateralization of the SLF. In the SLF, reported asymmetry varies in the literature, due in large part to whether the SLF was divided into sub-components and whether it was considered synonymous with the arcuate fasciculus.^20,71^ One current model divides the SLF into SLF I (which may actually be part of the cingulum),^72^ connecting the superior frontal and parietal cortices, SLF II, connecting superior and middle frontal gyri with the angular gyrus, SLF III, connecting the inferior frontal gyrus with the temporo-parietal junction, and the arcuate fasciculus, connecting the inferior frontal gyrus with the temporal cortex.^71,73^ The arcuate fasciculus is strongly left lateralized and may not even exist in the right hemisphere.^74^ The SLF III, however, shows a strong right lateralization.^20,50,72–74^ We did not divide the SLF into multiple components. Similarly to the cingulum findings above, exaggerated lateralization in the expected direction may indicate preservation of the dominant hemisphere at the expense of the non-dominant hemisphere. We did not have any harmonized measures of language function to test whether these alterations had functional consequences.

We also found that greater asymmetry in the uncinate fasciculus was associated with worse somatic symptoms. Studies of asymmetry in the UF have been mixed,^42,75,76^ although this may be because the laterality of the UF varies across tract segments.^77^ As we only considered the UF as a whole, we did not have the ability to identify asymmetry in the UF at a smaller scale. Follow-up analyses did not reveal any pattern in the shift in laterality, which may mean that injury leads to overall disorganization of the UF, or this may parallel prior studies with conflicting findings on the asymmetry of the UF as a whole.

Our study has several limitations. First, as a retrospective analysis of multi-site data collected in different studies, there are many sources of heterogeneity in our data that we cannot fully characterize or account for. Differences in the assessment of TBI history limited the analyses we could do to the most common denominators, which were often more general variables than individual sites collected. Our results present a number of interesting hypotheses that future studies can interrogate in greater detail in their datasets, such as whether the cingulum is vulnerable because it has an extended maturation time, whether females are less vulnerable to alterations in cingulum asymmetry because they have less natural cingulum asymmetry, or whether the right cingulum is more vulnerable to alterations due to less densely packed fibers. Second, most of the TBI history for our cohorts was self-reported, which has inherent issues with reliability. For this reason, we report all analyses of injury variables (such as duration of LOC) for completeness, but do not interpret these results. Third, TBSS is limited as a tensor-based approach when compared to a tractography approach, so we cannot fully attribute results to particular fiber bundles. Fourth, we were limited in the clinical endpoints we could examine to those that were most general (e.g., categorical variable for current depression), most commonly collected (e.g., TMT), or already harmonized (e.g., versions of the PCL). Work is ongoing in the ENIGMA Clinical Endpoints working group (a subgroup of the Brain Injury working group) to develop additional harmonized measures across multiple domains. Thus, future studies will be able to examine how TBI-related alterations in brain structure and function correspond to changes in memory, attention, and inhibition, among others. Fifth, as a cross-sectional study, we cannot examine whether asymmetry differences predate injury, or how the differences in asymmetry evolve over the lifespan. Lastly, while one cohort came from the Netherlands (BETTER/UMCU), the rest of the cohorts were from the United States, which may limit the generalizability of our results to other military contexts.

The effect sizes we report are small (Cohen’s *d*=0.18 for the main comparison), but in line with other ENIGMA analyses.^29,31^ Given the many sources of heterogeneity in our sample, and the existing, and often inconclusive, literature on dMRI alterations in military-relevant TBI (predominantly mTBI), small effect sizes are expected. The necessary sample size to find an effect of this size with 80% power and a significance threshold of 5% is 972, which is far larger than most published work on dMRI in military brain injury, highlighting the importance of collaborative projects, such as ENIGMA. Our work points to subtle alterations in the balance of the brain after TBI, and suggests a phenomenon of heightened vulnerability of the non-dominant hemisphere, perhaps as neural resources are diverted to support the dominant hemisphere. These alterations have functional consequences, as they were associated with slower processing speed and set shifting. Future targeted studies in individual, well-characterized cohorts or in animal models will aid in further elaborating on the specific mechanisms underlying, and on the functional implications for, alterations in tract asymmetry.

## Supporting information

STROBE

## Acknowledgements

R61NS120249; K99NS096116; This work was supported by U.S. Army Medical Research and Materiel Command (USAMRMC) award #13129004; VA RR&D IK2RX002922; VA R&D SPIRE Award; DOD PRARP; Dutch Ministry of Defence; R01AG058822; R01NS100973, DoD contract W81XWH-18-1-0413, a grant from the James J. and Sue Femino Foundation, a Hanson-Thorell Research Scholarship, the USC CURVE & SURE programs, and the Leonard Davis School of Gerontology; R01NS100952; European Research Council (ERC Starting Grant 804326); VA Rehabilitation Research and Development Service I01RX003443, I01RX003442; VA Health Services Research and Development Service Research Career Scientist Award IK6HX002608; This work was supported in part by Merit Review Award Number I01 CX001820 from the United States (U.S.) Department of Veterans Affairs Clinical Sciences R&D (CSRD) Service; VA CSR&D Career Development Award VA Career Development Award 5IK2CX001508; VA CSR&D Career Development Award 1IK2CX001772-01; Defense and Veterans Brain Injury Centers, the U.S. Army Medical Research and Materiel Command (USAMRMC; W81XWH-13-2-0025) and the Chronic Effects of Neurotrauma Consortium (CENC; PT108802-SC104835); U54EB020403; K99 MH119314 (NIMH); NARSAD 27786; I01 RX002174

