## Supplement for "Altered Lateralization of the Cingulum in Deployment-Related Traumatic Brain Injury: An ENIGMA Military-Relevant Brain Injury Study"

**Supplementary Information**

**Supplementary Note 1.** Further detail of ENIGMA-DTI protocols.

**Supplementary Table 1.** Clinical details and inclusion and exclusion criteria for each of the cohorts included.

**Supplementary Table 2.** Injury and comorbidity details collected across cohorts. For each cohort, we list the specific injury variables that were available. LOC=loss of consciousness, PTA=post-traumatic amnesia, PTSD=post-traumatic stress disorder, CES=Combat Exposure Scale, CAPS-IV/V=Clinician-Administered PTSD scale for DSM-IV/V, PCL=PTSD checklist (PCL-5=for DSM-V, PCL-C=for DSM-IV Civilian, PCL-M=for DSM-IV Military), DTS=Davidson Trauma Scale, GDS=Geriatric Depression Scale, SCID=Structured Clinical Interview for DSM-V, PHQ-9=Patient Health Questionnaire-9, CES-D=Center for Epidemiologic Studies – Depression, BDI-2=Beck Depression Inventory (BDI), SCL-90=Symptom Checklist-90, NSI=Neurobehavioral Symptom Inventory, BSI=Brief Symptom Inventory, DRRI-D=Deployment Risk and Resilience Inventory Section D.

**Supplementary Table 3.** DTI acquisition parameters for each of the cohorts included. For each cohort, we list the number of sites (for multi-site projects), the scanner manufacturers and models used, field strength (in Tesla, T), voxel size (in mm), number of gradient directions and the diffusion weighting (in $mm/s^2$), and the number of b0 volumes collected.

**Supplementary Note 2**. Symptom Inventories.

**Supplementary Table 4.** Supplementary group comparisons. dTBI=Deployment-related TBI. The analysis of participants whose worst injury was at age 45 or older did not include Vietnam Veterans as they are far more remote from their worst TBI. The black box outlining the cingulum results denotes the *post hoc* tests run, using a *p*-value of 0.05 to establish significance. The rest of the ROIs are listed for completeness. *T*-statistics and uncorrected *p*-values are shown, **bolded** *p*-values are those that survive correction for multiple comparisons, *italicized* *p*-values are those that do not survive multiple comparisons correction but were nominally significant (i.e., $0.05>p>0.003125$). ROI abbreviations may be found in **Supplementary Note 1.**

**Supplementary Table 5.** Interactions and associations with injury variables. *T*-statistics, unstandardized βs and uncorrected *p*-values are shown, *italicized* *p*-values are those that do not survive multiple comparisons correction ($0.05>p>0.003125$). ROI abbreviations may be found in **Supplementary Note 1.**

**Supplementary Figure 1.** Associations between cingulum laterality and Trail Making Task (TMT) performance. The two panels (left to right) show trend-level associations (with *p*-values) between TMT-A and TMT-B completion time (in seconds) and the cingulum laterality index (normalized residuals accounting for age, sex, and the nested random effects of cohort and site). Linear trend line shown with 95% confidence interval, calculated in R 3.6.0. Extreme outliers (>5 SD) were removed from the plots ($n$=4).

**Supplementary Note 1.** Further detail of ENIGMA-DTI protocols.

Details on scanner and acquisition parameters are provided in **Supplementary Table 3**. Preprocessing - including eddy current correction, echo planar imaging (EPI) induced distortion correction, and tensor fitting - was carried out at each site or at the central site for those sites that were able to share raw data. Image analysis was conducted using tract-based spatial statistics (TBSS) as part of the FSL software.[1] Individual subject fractional anisotropy (FA) maps were aligned to the custom ENIGMA-DTI FA template derived from 400 adult participants scanned across four sites designed for optimal multi-site harmonization.[2] FA voxels were then projected onto the ENIGMA-DTI template skeleton. This creates a unique FA skeleton in the same space for each individual in each cohort. To minimize effects of residual registration misalignment, the regions of interest were consistent in size across sites and the skeletonization procedure was performed individually at each site to minimize any site-specific residual misalignment. The same projection used for the FA images also projects the non-FA (mean, axial, and radial) images onto the skeleton. Voxels along the individual skeletons were averaged across white matter regions of interest (ROIs). A total of 25 bilateral ROIs were delineated based on the JHU white matter (WM) atlas, an established WM parcellation derived using deterministic tractography.[3] A whole-brain WM skeleton was defined according to the tract-based spatial statistics methodology [1] and ROI-averaged measures of FA, MD, AD and RD were then calculated by averaging each of these voxel measures over all skeleton voxels encapsulated by a particular ROI. This ensured that voxels at the periphery of a fiber bundle, where residual registration misalignment is typically maximal, were excluded from the ROI average. In other words, ROI averaging was performed based on the core of each fiber bundle, as defined by the WM skeleton.

The multi-subject JHU white matter parcellation atlas [3] was used to parcellate regions of interest from the ENIGMA template in MNI space, with updated label identification to correct an earlier atlas error.[4] A total of eighteen bilateral white matter ROIs were extracted from the skeletonized FA images and averaged (the corticospinal tract was ignored as prior reports have shown it to have poor reliability). The table below lists 24 ROIs (some partially overlapping) that were extracted from the skeletonized images, including 5 midsagittal regions (no lateralized components), and 18 lateralized regions (left and right are averaged to obtain bilateral FA). The overall average FA values were calculated by averaging values for the entire white matter skeleton.

ENIGMA-DTI QA/QC protocol consists of visual inspection of the images before and after registration to the ENIGMA template, as well as calculating the average skeleton projection distance. The distance of voxel projection to the ENIGMA skeleton can assess the registration quality between individual images and ENIGMA-DTI template. Higher projection distance may indicate problems with aligning individual brains to the template. After ROI extraction, histograms of FA and diffusivity measures are computed for each ROI.

| **Abbreviation** | **Full tract name** | **Abbreviation** | **Full tract name** |
|---|---|---|---|
| ACR (L+R) | Anterior *corona radiata* | PCR (L+R) | Posterior *corona radiata* |
| ALIC (L+R) | Anterior limb of internal capsule | PLIC (L+R) | Posterior limb of internal capsule |
| CGC (L+R) | Cingulum (cingulate gyrus) | PTR (L+R) | Posterior thalamic radiation |
| CGH (L+R) | Cingulum (hippocampal portion) | RLIC (L+R) | Retrolenticular part of internal capsule |
| CR (L+R) | *Corona radiata* | SCR (L+R) | Superior *corona radiata* |
| CST (L+R) | Corticospinal tract | SFO (L+R) | Superior fronto-occipital fasciculus |
| EC (L+R) | External capsule | SLF (L+R) | Superior longitudinal fasciculus |
| FXST (L+R) | *Fornix* (crus) / *Stria terminalis* | *SS (L+R)* | *Sagittal stratum* |
| IC (L+R) | Internal capsule | UNC (L+R) | *Uncinate* fasciculus |

| | | | |
|---|---|---|---|
| | | *TAP (L+R)* | *Tapetum* |

To examine potential group differences in subject motion, we extracted motion parameters from the *ecclog files generated by FSL eddy_correct. These were available for 8 of 16 cohorts, for a total of 1,692 participants, 749 with deployment-related TBI, 493 with civilian or undetermined TBI, and 450 controls. Following the approach in Ling et al.,[5] we examined rotation and translation, each averaged across the X, Y, and Z axes. We found no significant differences between the deployment-related TBI and control groups (rotation: $p$=0.087; translation: $p$=0.71). We note that these measures extracted by FSL's tool only quantify motion that is ultimately corrected for and may not fully capture motion-related artifacts left in the images. The ENIGMA-DTI protocol recommends sites perform visual inspection of the data at multiple processing steps to ensure images do not have severe artifacts, including those related to motion. The maximum skeletal projection distance is also quantified, and images of subjects with a maximal projection distance of 5 voxels are recommended for additional QC and removal.

**Supplementary Table 1.** Clinical details and inclusion and exclusion criteria for each of the cohorts included.

| Cohort | Inclusion criteria | Exclusion criteria | IRB |
|---|---|---|---|
| ADNI-DoD | All: Subjects must be Veterans of the Vietnam War, 50-90 years of age, must live within 150 miles of the closest ADNI clinic in subject's area. TBI: Subjects must have a documented history of moderate-severe non-penetrating TBI, which occurred during military service in Vietnam (identified from the Department of Defense or VA records). | All: MCI/Dementia, presence of PTSD by SCID-I for DSM-IV-TR criteria, or a CAPS score of >30 (Both current and/or a history of PTSD will be excluded). Control: Documented or self-report history of TBI, history of PTSD, MRI-related exclusions | Each of the 18 ADNI DoD sites' Institutional Review Boards as detailed in https://adni.loni.usc.edu/wp-content/uploads/2017/09/DODADNI_Procedures_Manual_20170912.pdf gave ethical approval for this work |
| BETTER/UMCU | All: 18-60 years of age, eligible for MRI. PTSD: current PTSD diagnosis, with CAPS ≥ 45, military deployment >4 months. Trauma controls: exposure to at least one traumatic event (according to DSM-IV A1 criterion), with CAPS < 15, no current psychiatric disorder, military deployment >4 months; healthy controls: no current psychiatric disorder according to DSM-IV. | All: history of neurological disorders, any severe or chronic disorder; alcohol or drug abuse and/or dependence during course of the study. | The Institutional Review Board for Utrecht University Medical Center gave ethical approval for this work |
| CENC | All: 1. deployment in Operation Enduring Freedom (OEF), Operation Iraqi Freedom (OIF), Operation New Dawn (OND), or follow-on conflicts, 2. history of combat exposure defined by the Deployment Risk and Resiliency Inventory Section D (DRRI-2-D) score >1 on any item, and 3. at least 18 years of age. | All: 1. any history of moderate/severe TBI as defined by either a) Glasgow Coma Scale >13, b) coma duration >0.5 hour, c) post-traumatic amnesia (PTA) duration >24 hours, or d) traumatic intracranial lesion; or 2. history of major neurologic disorder with significant decrease in functional status and/or loss of ability for independent living (e.g., dementia) or severe psychiatric disorder (e.g., schizophrenia). | The Institutional Review Boards at Hunter Holmes McGuire VA Medical Center, Michael E DeBakey VA Medical Center, James A Haley VA Hospital, South Texas Veterans Health Care System, Fort Belvoir Community Hospital, VA Portland Health Care System, Minneapolis VA Health Care System, and VA Boston Health Care System gave ethical approval for this work |
| Chronic Effects | All: 1. Age 18-62 years; 2. Right-handed; 3. Performs above cut-off scores on either the WMT or TOMM suggesting that effort is sufficient and patient's responses are valid; 4. Able to undergo MRI (e.g. no implanted metal, no embedded shrapnel, not pregnant); TBI: 1. Mild, moderate, or severe TBI received during OEF/OIF/OND deployment with at least one of the following:; a) Loss of Consciousness (LOC) ; b) Post-traumatic amnesia (PTA) ; c) Alteration of consciousness/mental state (AOC) (disorientation) >5 min; d) Worst post-resuscitation Glasgow Coma Scale (GCS) score 3-15 or Glasgow Coma Scale-Motor score 1-6 if intubated. (if GCS is available); 2. Post-injury interval > 2 months since most recent injury | All: 1. Not fluent in English; 2. Left-handed; 3. Pre-existing neurologic disorder associated with cerebral dysfunction and/or cognitive deficit; 4. Diagnosed learning disabilities or dyslexia. ; 5. Pre-existing severe psychiatric disorder (e.g., schizophrenia); 6. Pre-deployment moderate to severe TBI or post-deployment TBI with hospitalization; 7. Current alcohol or drug abuse assessed by AUDIT or DAST-10 on the day of brain imaging and neurobehavioral assessment; 8. Performs below cut-off on both the WMT and TOMM; 9. Contraindications to MRI; 10. Burn injury over more than 10% body area; 11. Unwilling to participate; TBI: 1. Coma, vegetative, minimally conscious state, or severe aphasia persists at time of attempted recruitment; 2. Non-evacuated or evacuated brain lesion >25 cc or midline shift >5 mm which could distort brain anatomy; Control: 1. Blast exposure; 2. Evidence of intracranial injury | The Institutional Review Board for Baylor College of Medicine and Affiliated Hospitals gave ethical approval for this work |

| | | | |
|---|---|---|---|
| Duke (both) | All: 18-65, OEF/OIF veterans, fluent in English, free of implanted metal objects or metal shards in eyes, antidepressant, sleep, and anti-anxiety medication permitted | All: Axis I other than PTSD or MDD, current substance abuse or lifetime substance dependence (other than nicotine), high risk for suicide, claustrophobia, neurological disorders, learning disability or developmental delay, major medical conditions | All: Axis I other than PTSD or MDD, current substance abuse or lifetime substance dependence (other than nicotine), high risk for suicide, claustrophobia, neurological disorders, learning disability or developmental delay, major medical conditions |
| HDFT | TBI Subjects: Age 18 to 70 years of either gender, documented history of TBI that either occurred in the last three months and required hospitalization (Acute) or occurred more than six months ago (Chronic), fluent in English. Control Subjects: Age 18 to 70 years of either gender, no prior history of concussion, TBI, blast exposure, stroke, or other major neurological disorder, fluent in English | Penetrating TBI due to gunshot wound, active drug or alcohol use or dependence that, in the opinion of the site investigator, would interfere with neuropsychological testing; inability or unwillingness of participant or legal guardian/representative (if the site has an IRB approved proxy consent) to provide written informed consent; contra-indication to MRI, such as ferrous metal, pacemakers, body weight above approximately 125 kg, or concerns about claustrophobia; history of any neurologic, psychiatric, or developmental disorders that, in the opinion of the investigator, may affect neuropsychological testing. | The Institutional Review Board for Baylor College of Medicine and Affiliated Hospitals gave ethical approval for this work |
| Houston Blast | All: 1. OEF/OIF/OND deployment; 2. Age 18 – 65; 3. Post-injury interval > 3 months ; 4. Use of at least one upper extremity; 5. Accepted by MRI technologist and neuroradiologist as safe to undergo MRI ; 6. Performs above cut-off scores on the Word Memory Task (WMT) (> 82.5%) and Test of Memory Malingering (TOMM) (> 40 on Trial 1 and > 44 on Trial 2) [53].; 7. Readiness for fMRI (i.e., can be trained to perform cognitive tasks at criterion level prior to scanning, intact visual fields, mild or no visual neglect, and adequate visual acuity); TBI: 1. Mild deployment-related TBI | All: 1. Pre-existing neurologic disorder associated with cerebral dysfunction and/or cognitive deficit, diagnosed learning disability or dyslexia; 2. Current severe alcohol (AUDIT>15) or drug abuse (DAST-10 >3) or pre-existing severe psychiatric disorder (e.g., schizophrenia or bipolar disorder ); 3. Performs below cut-off on one or both the WMT and TOMM; 4. Unable to use at least one upper extremity; 5. Contraindications to MRI and fMRI; 6. Unwilling to participate; TBI: 1. Moderate to severe deployment-related TBI; 2. History of a trauma event resulting only in disorientation for < 5 minutes, but no LOC or PTA; 3. History of pre -deployment TBI that required hospitalization; 4. History of post-deployment TBI or concussion; Control: 1. history of pre- or post-deployment TBI or concussion; 2. evidence of intracranial injury | The Institutional Review Board for Baylor College of Medicine and Affiliated Hospitals gave ethical approval for this work |
| INTRuST | For complete inclusion criteria see (Bomyea et al. 2019) | For complete exclusion criteria see (Bomyea et al. 2019) | The INTRuST IRB gave ethical approval for this work |
| iSCORE | TBI: presence of persistent cognitive symptoms as measured by a Neurobehavioral Symptom Inventory (NSI) score of 3 or higher on any of the four cognitive symptoms, ages 18 to 55 years, sustained a closed head injury during deployment (OEF/OIF/OND) activities 3 to 24 months prior to recruitment, English proficiency, and no PTSD diagnosis. PTSD: met clinical criterion for a PTSD diagnosis based on the CAPS due to combat related trauma only, no previous history of closed head injury, age 18 to 55, deployment history 3 to 36 months, and English proficiency. Deployment history inclusion criteria had to be extended by 12 months in this particular medical setting as PTSD-associated symptoms requiring intensive treatment were often identified and treated long after being deployed. Control: no previous history of closed | Any participants with conditions preventing MRI procedures (i.e. claustrophobia, shrapnel, pregnancy), neurologic conditions (e.g., seizures, psychosis, etc.), history of TBI exceeding mild severity or spinal cord injury, or current narcotic medicine use were excluded from the study. | The Brooke Army Medical Center Institutional Review Board gave ethical approval for this work |

| | | | |
|---|---|---|---|
| | head injury, no PTSD diagnosis based on the CAPS, age 18 to 55, deployment history 3 to 24 months, and English proficiency. | | |
| NICoE | 1. History or evaluation of head trauma or post-concussive symptoms; 2. Active duty or DEERS eligible individuals (to include non-active duty, military healthcare beneficiaries); 3. Adult between the ages of 18 and 60; 4. Males and non-pregnant/non-breastfeeding females (due to neuroimaging). | 1. Traumatic brain injury patients who are unable to consent themselves; 2. Actively enrolled in other randomized controlled treatment trials where this study would interfere; 3. History of prior severe neurologic or psychiatric condition, such as psychosis, stroke, multiple sclerosis, or spinal cord injury; 4. Pregnancy (by history and urine assay); 5. Breastfeeding. | The Institutional Review Board for the Walter Reed National Military Medical Center gave ethical approval for this work |
| Rover Wiser | All: Age 18-50 years, Right-handed (as determined by the Edinburgh Handedness Inventory by a score > 40), Able to safely and comfortably undergo MRI (e.g. no implanted metal, no embedded shrapnel, claustrophobia, etc.), participants are required to have been previously deployed in OEF/OIF/OND. TBI: History of mTBI based on at least one of the following - LOS < 24 hours, PTA > 7 days, AOC 5min-24hrs, GCS=9-15 (if available). Control: Uninjured, or sustaining a non-cranial injury without history of blast exposure, Post-injury interval > 3 months or more since most recent injury (for the participants with non-cranial injuries) | All: Not fluent in English, Ever having been formally diagnosed with dyslexia or other learning disabilities, Left-handed (as determined by the Edinburgh Handedness Inventory by a score < 40), Post-deployment TBI requiring hospitalization (i.e., not just treated and released directly from Emergency Center ), Contraindications to undergoing MR imaging (e.g., metal implants, orthodontia, shrapnel, positive urine pregnancy screen, claustrophobia, etc.), Pre- or post-deployment neurologic disorder associated with cerebral dysfunction and/or cognitive deficit (e.g., mental retardation, HIV/AIDS, dementias, etc.), Pre-deployment major psychiatric disorder associated with cerebral dysfunction and/or cognitive deficit (e.g., schizophrenia and other psychotic disorders, bipolar disorder),as determined by the Structured Clinical Interview for the DSM-V conducted by ROVER-WISER clinicians (or the most current version of the MINI for Veteran Controls—see criteria for Controls below), Current active psychosis, Any history of intracranial surgery, Medical conditions associated with structural/functional compromise on MRI or cognitive decrements; Control: 1. Meets MINI criteria for any substance use disorders (SUDs) other than nicotine (tobacco products), Meets MINI criteria for current depressive episode, Meets MINI criteria for PTSD, Meets MINI criteria for other major psychiatric disorder associated with cerebral dysfunction and/or cognitive deficit (e.g., schizophrenia and other psychotic disorders, bipolar disorder), Meets criteria for history of TBI of any severity | The Institutional Review Board for Baylor College of Medicine and Affiliated Hospitals gave ethical approval for this work |
| SPIRE/GBET | Inclusion criteria. All subjects will meet the following inclusion criteria: 1. Meet criteria for mTBI as defined by the VA/DoD Clinical Practice Guideline. 2. Meet criteria for PTSD as assessed by the Clinician Administered PTSD Scale (CAPS; Blake et al., 1995); 3. Aged 18 - 49; 4. Male; 5. Have been referred for treatment by a clinician in the PTSD Clinical Team (PCT) at the Michael E. DeBakey VA medical Center (MEDVAMC); 6. Have not previously participated in meditation training, and 7. Are cleared to participate by a treating MEDVAMC clinician. | Exclusion criteria. We will exclude subjects who: 1. Meet DSM-IV criteria for drug or alcohol abuse in past 30 days; 2. Have a history of severe TBI based on any of following: (i) Glasgow Coma Score < 8; (ii) alteration of consciousness > 24 hours; loss of consciousness > 30 minutes; 3. Have current neurological or general medical conditions known to impact cognitive and/or emotional functioning, including but not limited to: epilepsy, Parkinson's disease, Huntington's disease, Alzheimer's disease, stroke, chemotherapy for cancer; 4. Have acute psychological instability as assessed by MEDVAMC clinician or study staff or concurrent diagnosis or schizophrenia, schizoaffective disorder, delusional disorder, organic psychosis, and subjects taking antipsychotic medication, and 5. Have already completed a course of meditation training. We will also exclude participants with general contraindications for MRI, including | The Institutional Review Board for Baylor College of Medicine and Affiliated Hospitals gave ethical approval for this work |

| | | | |
|---|---|---|---|
| | | metal in or around the head (e.g., orthodontia, non-removable body piercings, etc.), ferromagnetic material in the body (e.g., non-removable body piercings), or non-MRI compatible medical devices. | |
| VA Minneapolis (both) | Participants were veterans of Operation Enduring Freedom and/or Operation Iraqi Freedom, age 22-60, who had been exposed to combat during their deployment(s). | Participants were excluded from the study if they met criteria for 1. a current substance-induced psychotic disorder or psychotic disorder due to a general medical condition (other than TBI), 2. current DSM-IV substance abuse or dependence other than alcohol, caffeine, or nicotine, 3. a moderate or severe traumatic brain injury from either impact or blast, 4. a neurologic condition other than TBI, 5. a current unstable medical condition that would likely affect brain function (e.g., uncontrolled diabetes), or 6. significant imminent risk of suicidal or homicidal behavior. | The Minneapolis VA Health Care System Institutional Review Board gave ethical approval for this work |
| VETSA | Participants were recruited from the Vietnam Era Twin Registry aka VETR (by registry definition, then, both brothers had been in the US military at some point between 1965 and 1975; not "VA"). Participants had to be between 50 to 59 years old when recruited; both brothers needed to agree to participate. | For MRI component, participants needed to meet safety criteria. | The Institutional Review Boards for Boston University and the University of California, San Diego gave ethical approval for this work |

**Supplementary Table 2.** Injury and comorbidity details collected across cohorts. For each cohort, we list the specific injury variables that were available. LOC=loss of consciousness, PTA=post-traumatic amnesia, PTSD=post-traumatic stress disorder, CES=Combat Exposure Scale, CAPS-IV/V=Clinician-Administered PTSD scale for DSM-IV/V, PCL=PTSD checklist (PCL-5=for DSM-V, PCL-C=for DSM-IV Civilian, PCL-M=for DSM-IV Military), DTS=Davidson Trauma Scale, GDS=Geriatric Depression Scale, SCID=Structured Clinical Interview for DSM-V, PHQ-9=Patient Health Questionnaire-9, CES-D=Center for Epidemiologic Studies – Depression, BDI-2=Beck Depression Inventory (BDI), SCL-90=Symptom Checklist-90, NSI=Neurobehavioral Symptom Inventory, BSI=Brief Symptom Inventory, DRRI-D=Deployment Risk and Resilience Inventory Section D.

| Cohort | Injury context | Blast | Mechanism | Multiple injuries queried | Num. TBI reported | TSI Reported | Chronicity | LOC/PTA | PTSD scale | Depression scale | Symptom scale | CES (test: mean, SD, range) |
|---|---|---|---|---|---|---|---|---|---|---|---|---|
| **ADNIDoD** | Non-military /deployment | No | No | Yes | Yes | Yes - first, worst, recent | Decades | Yes | CAPS-IV | GDS | SCL-90 | CES: 17.7, 11.4, 0-41 |
| **BETTER /UMCU** | Deployment | Yes | Yes | No | No | No | NA | Yes | CAPS-IV | SCID | NA | NA |
| **CENC** | Non-military /deployment | Yes | Yes | Yes | Yes | Yes - first, recent | >1 year | Yes | PCL-5 | PHQ-9 | NSI | DRRI-D: 37.4, 15.2, 17-93 |
| **Chronic Effects** | Deployment | Yes | Yes | Yes | Yes - blast | Yes - recent, worst | >1 year | Yes | PCL-C | CES-D | NSI | CES: 12.7, 8.0, 0-29 |
| **Duke1** | Non-military /deployment | Yes | Yes | Yes | No | Yes - recent, worst | >1 year | Yes - LOC only | CAPS-IV/V | BDI-2 | BSI | NA |
| **Duke2** | Non-military /deployment | Yes | Yes | Yes | No | No | NA | Yes - LOC only | DTS/SCID | NA | SCL-90 | CES: 11.6, 10.5, 0-35 |
| **HDFT** | Non-military /deployment | Yes | Yes | No | Yes | Yes - recent, worst | >6 months | Yes | PCL-C | NA | NA | NA |
| **Houston Blast** | Deployment | Yes | Yes | Yes | No | Yes - recent, worst | 1-10 years | Yes | PCL-C | CES-D | NSI (34 item) | NA |
| **INTRuST** | No | Yes | Yes | No | No | No | NA | Not collected | PCL-M | NA | NA | NA |
| **iSCORE** | All combat-related | Yes | Yes | No | Yes | Yes - recent, worst | 2 months - 2 years | Yes | PCL-M | CES-D | NSI | NA |
| **NICoE** | Non-military /deployment | Yes | Yes | Yes | Yes | Yes - first, worst, recent | 2 months - 40 years | Yes | PCL-C | NA | NA | NA |
| **Rover Wiser** | Non-military /deployment | Yes | Yes | Yes | Yes - blast | Yes - recent, worst | >4 months | Yes | CAPS-IV | BDI-2 | NA | NA |
| **SPIRE/GBET** | Non-military /deployment | Yes | No | No | No | No | NA | No | CAPS-IV | BDI-2 | NA | DRRI-D: 46.2, 18.1, 17-90 |
| **VA Minneapolis1** | Non-military /deployment | Yes | Yes | Yes | Yes | Yes - recent, worst | >3 months | Yes - no duration | CAPS-IV | SCID | NA | CES: 19.7, 10.8, 0-41 |
| **VA Minneapolis2** | Non-military /deployment | Yes | Yes | Yes | Yes | Yes - recent, worst | >8 months | Yes - no duration | CAPS-IV | SCID | NA | NA |
| **VETSA** | Non-military /deployment | No | Yes | Yes | Yes | Yes - first, worst, recent | Decades | Yes | PCL-C | CES-D | NA | NA |

**Supplementary Table 3.** DTI acquisition parameters for each of the cohorts included. For each cohort, we list the number of sites (for multi-site projects), the scanner manufacturers and models used, field strength (in Tesla, T), voxel size (in mm), number of gradient directions and the diffusion weighting (in $mm/s^2$), and the number of b0 volumes collected.

| Cohort | Number of sites | Scanner | Field strength | Voxel size (mm) | Gradient directions and b-value ($mm/s^2$) | No. b0 volumes |
|---|---|---|---|---|---|---|
| **ADNIDoD** | 15 | 1. GE Discovery MR750, 2. GE Discovery MR750w, 3. GE Signa HDxt, 4. Siemens TrioTim | 3T | 2x2x2 | 41 at b=1000 | 5 |
| **BETTER/UMCU** | 1 | Philips Achieva | 3T | 1.875x1.875x2 | 30 at b=1000 | 1 |
| **CENC** | 8 | 1. Philips Ingenia, 2. Siemens TrioTim, 3. GE Signa HDxt, 4. Siemens Verio/Skyra Fit, 5. GE Discovery MR750, 6. Philips Achieva, 7. Siemens Prisma, 8. Siemens Prisma | 3T | Sites 1,2,4,6: 2.7 iso, 3. 1.3x1.3x2.7, 5. 1.4x1.4x2.7, Sites 7 and 8: 1.5 iso | Sites 1, 2, 4, 5: 64 at b=1300, 3. 60 at b=1300, 6. 32 at b=1000, Sites 7 and 8: 140 at b=1000 and 140 at b=2000 | Sites 1-6: 8, Site 7 and 8: 38 |
| **Chronic Effects** | 1 | Siemens TrioTim | 3T | 2.7x2.7x2.5 | 30 at b=1000 | 1 |
| **Duke1** | 1 | GE Discovery MR750 | 3T | 2x2x2 | 64 at b=900 | 5 |
| **Duke2** | 1 | Philips Ingenia | 3T | 2x2x2 | 32 at b=800 | 2 |
| **HDFT** | 1 | Siemens TrioTim | 3T | 2.4x2.4x2.4 | 65 at b=1000 | 12 |
| **Houston Blast** | 1 | Siemens TrioTim | 3T | 2.7x2.7x2.5 | 30 at b=1000 | 1 |
| **INTRuST** | 11 | Philips | 3T | 2x2x2 | 87 at b=900 | 7 |
| **iSCORE** | 1 | Siemens Verio Syngo | 3T | 2x2x2 | 64 at b=1000 | 1 |
| **NICoE** | 1 | GE Discovery MR750 | 3T | 1.7x1.7x1.7 | 90 at b=1000 | 19 |
| **Rover Wiser** | 1 | Siemens Trio | 3T | 2.7x2.7x3.2 | 30 at b=1000 | 1 |
| **SPIRE/GBET** | 1 | Siemens Trio | 3T | 2.7x2.7x3.2 | 30 at b=1000 | 1 |
| **VA Minneapolis1** | 1 | Siemens TrioTim | 3T | 2x2x2 | 30 at b=800 | 10 |
| **VA Minneapolis2** | 1 | Siemens TrioTim | 3T | 2x2x2 | 128 at b=1500 | 17 |
| **VETSA** | 1 | 1. GE Discovery MR750, 2. Siemens TrioTim | 3T | 2.5x2.5x2.5 | 1. 51 at b=1000, 2. 30 at b=1000 | 1. 2, 2. 1 |

**Supplementary Note 2**. Symptom Inventories.

As depression was assessed using a range of scales, we used published clinical cutoffs to establish a categorical depression variable. For the Beck Depression Inventory (BDI), the cutoff for depression was >13.[7] For the Geriatric Depression Scale (GDS), the cutoff for depression was >4.[8] For the Center for Epidemiologic Studies – Depression scale (CES-D), the cutoff for depression was >15.[9] For the Patient Health Questionnaire-9 (PHQ-9), the cutoff for depression was >. The Structured Clinical Interview for DSM-V (SCID) yields a categorical variable not a score so it did not need to be converted.

For posttraumatic stress disorder (PTSD), the most commonly collected symptom measure was the PTSD Checklist (PCL). There was variety in the version collected, with some collecting the PCL-5 (based on DSM-V) and others collecting the PCL-4 (based on DMV-IV) which is divided into a Civilian version (PCL-C), and a Military version (PCL-M). As part of the harmonization process, PCL-4 scores were converted to PCL-5 scores, and for the PCL, the cutoff for PTSD was >33.[10,11] Other PTSD measures collected include the Clinician-Administered PTSD Scale for DSM-IV/V (CAPS-IV/V), for which the cutoff was >40,[12] the Davidson Trauma Scale using a cutoff of >40,[13] or the SCID. The equation for converting PCL-C/M scores to PCL-5, as reported in,[14] is:

$$PCL5 = -33.5 + 1.82(PCLC/M) - 0.0065(PCLC/M)^2$$

Seven sites collected the Symptom Checklist 90 (SCL-90)[15], Brief Symptom Inventory (BSI)[16], or Neurobehavioral Symptom Inventory (NSI - 22 or 34 item version).[17] Prior psychometric analyses on the factor structure of these inventories have shown that several common factors can be extracted from these: somatization and affective.[18–22] The SCL-90 and BSI further have the affective factor separated into depression and anxiety, while the NSI does not. For this analysis, we generated somatization and affective scores from the item level data across seven sites, summing across the items belonging to each factor and dividing by the highest total possible, yielding a 0-1 measure for each symptom domain.

**Supplementary Table 4.** Supplementary group comparisons. dTBI=Deployment-related TBI. The analysis of participants whose worst injury was at age 45 or older did not include Vietnam Veterans as they are far more remote from their worst TBI. The black box outlining the cingulum results denotes the *post hoc* tests run, using a *p*-value of 0.05 to establish significance. The rest of the ROIs are listed for completeness. *T*-statistics and uncorrected *p*-values are shown, **bolded** *p*-values are those that survive correction for multiple comparisons, *italicized* *p*-values are those that do not survive multiple comparisons correction but were nominally significant (i.e., 0.05>*p*>0.003125). ROI abbreviations may be found in **Supplementary Note 1.**

| | dTBI (under 40 years old, *n*=1,647) | dTBI (over 45 years old, *n*=898) | dTBI (males, *n*=1,678) | dTBI (females, *n*=194) | dTBI (cov. current depression, *n*=1,653) | dTBI (cov. current PTSD, *n*=1,840) | dTBI (cov. current depression and PTSD, *n*=1,645) | dTBI (cov. for handedness, *n*=1,352) | dTBI (only right handed participants, *n*=1,183) | dTBI (only Vietnam Veterans, *n*=253) | dTBI (only OEF/OIF/OND ADSMV, *n*=1,619) |
|---|---|---|---|---|---|---|---|---|---|---|---|
| **ACR** | -0.61 (0.54) | 1.2 (0.22) | -1.1 (0.26) | 1.4 (0.16) | -0.088 (0.93) | -0.48 (0.63) | -0.23 (0.82) | 0.64 (0.52) | 0.73 (0.46) | 0.15 (0.88) | -0.31 (0.76) |
| **ALIC** | -0.078 (0.94) | -0.80 (0.42) | 0.76 (0.45) | 0.69 (0.49) | 1.2 (0.23) | 0.80 (0.42) | 1.1 (0.27) | 1.1 (0.29) | 0.79 (0.43) | 1.2 (0.24) | 0.78 (0.44) |
| **CGC** | **2.4 (0.017)** | 0.21 (0.83) | **3.4 (0.00072)** | 1.8 (0.077) | **3.3 (0.0011)** | **3.4 (0.00058)** | **3.1 (0.0022)** | **3.7 (0.00020)** | **4.1 (0.000053)** | 2.0 (0.051) | **3.5 (0.00047)** |
| **CGH** | 0.14 (0.89) | -0.15 (0.88) | -0.17 (0.87) | 1.4 (0.15) | 0.16 (0.88) | -0.082 (0.93) | -0.058 (0.95) | 0.55 (0.58) | 0.13 (0.90) | 0.56 (0.57) | 0.17 (0.86) |
| **CST** | -1.3 (0.19) | -1.5 (0.14) | -1.6 (0.12) | -1.1 (0.26) | -1.4 (0.17) | -1.6 (0.10) | -1.4 (0.16) | -1.2 (0.23) | -0.66 (0.51) | -1.8 (0.069) | -1.2 (0.23) |
| **CR** | 0.064 (0.95) | *2.4 (0.019)* | 1.2 (0.22) | 0.52 (0.61) | 1.2 (0.23) | 1.6 (0.11) | 1.3 (0.20) | 0.61 (0.54) | 0.29 (0.77) | 0.25 (0.80) | 1.2 (0.22) |
| **EC** | -1.1 (0.27) | 1.0 (0.33) | -0.40 (0.69) | *-2.1 (0.035)* | 0.56 (0.58) | -0.56 (0.57) | 0.29 (0.77) | -1.2 (0.23) | -0.95 (0.34) | -0.70 (0.48) | -0.49 (0.62) |
| **FX/ST** | -0.086 (0.93) | 0.93 (0.35) | 0.81 (0.42) | -0.35 (0.73) | 0.25 (0.80) | 0.18 (0.86) | -0.039 (0.97) | 1.2 (0.22) | 1.3 (0.20) | 0.12 (0.91) | 0.69 (0.49) |
| **IC** | -0.80 (0.42) | 0.81 (0.42) | 0.81 (0.42) | -1.2 (0.23) | 0.64 (0.52) | 0.28 (0.78) | 0.62 (0.53) | 0.51 (0.61) | 0.68 (0.50) | -1.5 (0.14) | 0.69 (0.49) |
| **PCR** | 0.55 (0.59) | -0.51 (0.61) | 1.0 (0.32) | *-0.63 (0.53)* | 1.0 (0.31) | 0.64 (0.52) | 0.83 (0.41) | 0.27 (0.79) | 0.42 (0.67) | 1.4 (0.15) | 0.56 (0.58) |
| **PLIC** | -0.64 (0.52) | 0.68 (0.49) | 0.53 (0.60) | -1.3 (0.18) | -0.050 (0.96) | -0.081 (0.94) | -0.11 (0.91) | -0.05 (0.96) | 0.41 (0.68) | -1.6 (0.10) | 0.37 (0.71) |
| **PTR** | 0.63 (0.53) | 1.1 (0.28) | 1.0 (0.30) | 1.5 (0.14) | 1.6 (0.10) | 1.2 (0.21) | 1.4 (0.15) | 1.5 (0.14) | 0.76 (0.45) | 1.3 (0.19) | 1.4 (0.16) |
| **RLIC** | -0.46 (0.65) | 1.0 (0.30) | -0.0024 (1.00) | -0.08 (0.94) | 0.90 (0.37) | 0.32 (0.75) | 0.91 (0.36) | 0.21 (0.84) | -0.080 (0.94) | -1.4 (0.17) | 0.11 (0.91) |
| **SCR** | 1.2 (0.24) | -0.93 (0.35) | 1.6 (0.10) | -1.2 (0.25) | 1.5 (0.14) | 1.7 (0.091) | 1.6 (0.10) | 0.97 (0.33) | 1.2 (0.22) | -0.23 (0.82) | 1.5 (0.14) |
| **SFO** | -0.34 (0.73) | 0.031 (0.98) | -1.1 (0.28) | 0.73 (0.47) | -0.25 (0.80) | -0.21 (0.83) | -0.053 (0.96) | -0.53 (0.60) | -0.51 (0.61) | -1.2 (0.24) | -0.23 (0.82) |
| **SLF** | 1.1 (0.29) | 2.2 (0.027) | 2.4 (0.015) | -0.39 (0.70) | 2.8 (0.0056) | 2.4 (0.016) | 2.8 (0.0060) | 2.2 (0.026) | 2.5 (0.011) | **3.0 (0.0030)** | 1.5 (0.12) |
| **SS** | 0.63 (0.53) | *2.8 (0.0056)* | *1.8 (0.071)* | -0.32 (0.75) | *1.0 (0.30)* | *0.84 (0.40)* | *0.75 (0.45)* | *1.6 (0.12)* | *0.86 (0.39)* | 0.26 (0.79) | 1.4 (0.15) |
| **TAP** | -1.1 (0.29) | *-2.1 (0.035)* | -1.6 (0.11) | -1.4 (0.16) | -0.96 (0.34) | -1.6 (0.11) | -1.2 (0.23) | -1.3 (0.18) | -0.79 (0.43) | -1.9 (0.06) | -1.4 (0.16) |
| **UNC** | -0.11 (0.91) | *0.76 (0.45)* | 0.38 (0.71) | 2.3 (0.022) | 0.98 (0.33) | 0.90 (0.37) | 0.93 (0.35) | 0.90 (0.37) | 0.75 (0.45) | -0.082 (0.93) | 1.2 (0.23) |

**Supplementary Table 5.** Interactions and associations with injury variables. *T*-statistics, unstandardized βs and uncorrected *p*-values are shown, *italicized p*-values are those that do not survive multiple comparisons correction (0.05>*p*>0.003125). ROI abbreviations can be found in **Supplementary Note 1.**

| | Group-by-Age | Group-by-Gender | HX of TBI with PTA | Time since first injury | Time since worst injury | Time since most recent injury | Number of TBI | Number of blast-related TBI | Length of LOC | Length of PTA | Number of TBI with LOC | Number of TBI with PTA |
|---|---|---|---|---|---|---|---|---|---|---|---|---|
| **ACR** | 4.8e-5 (0.74) | 0.012 (0.029) | 0.49 (0.62) | -1.2e-5 (0.82) | -5.3e-5 (0.46) | -3.7e-5 (0.63) | 5.2e-4 (0.18) | 2.4e-4 (0.80) | 9.6e-4 (0.54) | 2.1e-3 (0.37) | 1.1e-3 (0.11) | 1.2e-3 (0.078) |
| **ALIC** | -3.0e-4 (0.16) | 0.0033 (0.68) | 0.13 (0.90) | 3.3e-5 (0.58) | 3.8e-5 (0.65) | -1.0e-5 (0.91) | 6.1e-4 (0.17) | -1.1e-4 (0.92) | 9.7e-4 (0.61) | 6.2e-4 (0.79) | 1.0e-3 (0.18) | 1.3e-3 (0.079) |
| **CGC** | -1.3e-4 (0.67) | 0.0086 (0.46) | 1.9 (0.055) | 3.0e-5 (0.81) | 4.5e-5 (0.76) | 7.6e-5 (0.62) | -4.3e-4 (0.64) | -7.5e-4 (0.71) | -2.9e-3 (0.35) | 3.8e-3 (0.33) | 1.5e-3 (0.33) | 2.1e-3 (0.15) |
| **CGH** | 7.4e-5 (0.76) | 0.010 (0.26) | 1.3 (0.19) | -4.8e-5 (0.62) | -3.5e-5 (0.78) | -8.0e-5 (0.54) | 5.1e-4 (0.48) | -1.1e-3 (0.52) | 2.7e-3 (0.23) | *8.7e-3 (0.022)* | -6.5e-4 (0.60) | 7.8e-5 (0.96) |
| **CST** | *-2.2e-4 (0.0093)* | -0.0013 (0.68) | -1.0 (0.30) | -4.4e-5 (0.15) | -4.4e-5 (0.29) | -5.1e-5 (0.25) | 1.7e-5 (0.94) | -2.5e-4 (0.65) | 7.0e-4 (0.38) | -6.7e-4 (0.58) | 4.0e-4 (0.30) | 6.3e-4 (0.13) |
| **CR** | 1.3e-4 (0.60) | 0.0015 (0.87) | 1.6 (0.11) | 1.8e-5 (0.85) | 2.6e-5 (0.84) | -1.3e-6 (0.99) | 2.6e-4 (0.71) | -9.7e-4 (0.52) | 4.2e-3 (0.093) | -3.5e-3 (0.35) | *2.5e-3 (0.034)* | *3.6e-3 (0.0061)* |
| **EC** | -1.7e-4 (0.29) | -0.0067 (0.27) | 0.51 (0.61) | -5.7e-5 (0.30) | -5.1e-5 (0.50) | -5.4e-6 (0.94) | -3.2e-4 (0.45) | -1.3e-4 (0.88) | -2.4e-3 (0.13) | -4.7e-4 (0.81) | -3.8e-4 (0.58) | 4.1e-4 (0.56) |
| **FX/ST** | -7.1e-5 (0.72) | -0.0049 (0.51) | -0.43 (0.67) | -2.9e-5 (0.68) | -3.5e-5 (0.73) | 1.2e-5 (0.91) | 7.8e-5 (0.88) | -1.6e-3 (0.23) | 3.5e-3 (0.083) | -1.9e-3 (0.56) | 3.4e-4 (0.71) | -9.0e-4 (0.36) |
| **IC** | -9.9e-5 (0.35) | -0.0038 (0.33) | -0.24 (0.81) | -3.2e-5 (0.37) | -2.0e-5 (0.69) | *-1.1e-4 (0.041)* | -1.4e-4 (0.59) | 1.7e-4 (0.77) | 1.4e-3 (0.16) | -1.1e-3 (0.42) | 1.4e-4 (0.76) | 4.0e-4 (0.39) |
| **PCR** | -2.5e-5 (0.86) | -0.0032 (0.54) | 0.38 (0.71) | *1.0e-4 (0.049)* | 1.2e-4 (0.069) | *1.5e-4 (0.034)* | 4.6e-4 (0.21) | 1.6e-4 (0.86) | 7.5e-4 (0.58) | *-4.4e-3 (0.036)* | 5.9e-4 (0.37) | 1.2e-3 (0.093) |
| **PLIC** | -1.2e-4 (0.51) | -0.0090 (0.16) | -1.6 (0.12) | -1.0e-4 (0.067) | -7.3e-5 (0.39) | -1.4e-4 (0.11) | -6.1e-4 (0.17) | 3.0e-5 (0.97) | 7.0e-4 (0.69) | -3.6e-4 (0.85) | -1.2e-3 (0.081) | -8.6e-4 (0.23) |
| **PTR** | 6.1e-5 (0.75) | 0.0055 (0.44) | 0.40 (0.69) | 2.4e-5 (0.68) | 8.7e-5 (0.29) | 8.5e-5 (0.33) | 3.6e-5 (0.93) | -4.7e-4 (0.66) | 3.3e-4 (0.85) | -4.7e-3 (0.062) | 5.9e-4 (0.43) | 9.8e-4 (0.23) |
| **RLIC** | 7.3e-5 (0.61) | 0.0018 (0.74) | 0.46 (0.65) | -2.9e-5 (0.60) | -5.9e-5 (0.40) | -6.2e-5 (0.39) | -4.9e-5 (0.90) | 1.0e-4 (0.91) | -2.1e-3 (0.15) | -2.1e-3 (0.34) | 7.3e-4 (0.27) | 9.7e-4 (0.19) |
| **SCR** | *-2.7e-4 (0.010)* | -0.0066 (0.10) | 1.3 (0.19) | 1.7e-5 (0.68) | 5.3e-5 (0.32) | 2.6e-5 (0.65) | 2.3e-5 (0.94) | 1.9e-4 (0.80) | 1.9e-3 (0.069) | *4.3e-3 (0.0093)* | 6.9e-5 (0.89) | 7.0e-4 (0.23) |
| **SFO** | -1.3e-4 (0.64) | 0.011 (0.31) | -0.87 (0.38) | -1.3e-4 (0.19) | *-2.9e-4 (0.022)* | *-2.9e-4 (0.033)* | 3.4e-4 (0.65) | 2.2e-3 (0.24) | 7.0e-4 (0.79) | -4.3e-3 (0.19) | -4.1e-4 (0.75) | -4.0e-4 (0.74) |
| **SLF** | *3.7e-4 (0.0032)* | -0.0033 (0.49) | 1.9 (0.061) | *1.2e-4 (0.016)* | *1.4e-4 (0.025)* | *1.6e-4 (0.018)* | 2.9e-4 (0.42) | 5.3e-4 (0.52) | 2.6e-3 (0.058) | *5.3e-3 (0.0076)* | 1.2e-3 (0.054) | 9.1e-4 (0.19) |
| **SS** | 1.2e-4 (0.54) | -0.0049 (0.52) | -0.37 (0.71) | 2.0e-6 (0.98) | 5.6e-5 (0.59) | -4.8e-5 (0.65) | -3.1e-4 (0.57) | 3.4e-4 (0.79) | 2.1e-3 (0.34) | 1.1e-3 (0.70) | 5.0e-5 (0.95) | -7.7e-4 (0.32) |
| **TAP** | *-8.8e-4 (0.010)* | 0.00015 (0.99) | -0.75 (0.45) | -1.8e-4 (0.10) | -1.9e-4 (0.18) | -1.6e-4 (0.30) | -8.9e-4 (0.27) | -3.1e-5 (0.99) | -2.2e-3 (0.50) | -6.9e-3 (0.11) | -6.4e-4 (0.65) | 1.3e-4 (0.93) |
| **UNC** | 4.3e-5 (0.92) | *0.033 (0.045)* | 0.25 (0.80) | *-2.8e-4 (0.047)* | -2.1e-4 (0.28) | -1.9e-4 (0.34) | 1.2e-4 (0.91) | 3.1e-3 (0.18) | 7.7e-3 (0.046) | 4.6e-4 (0.93) | 2.0e-3 (0.25) | 2.3e-3 (0.21) |

**Supplementary Figure 1.** Associations between cingulum laterality and Trail Making Task (TMT) performance. The two panels (left to right) show trend-level associations (with *p*-values) between TMT-A and TMT-B completion time (in seconds) and the cingulum laterality index (normalized residuals accounting for age, sex, and the nested random effects of cohort and site). Linear trend line shown with 95% confidence interval, calculated in R 3.6.0. Extreme outliers (>5SD) were removed from the plots (*n*=4).

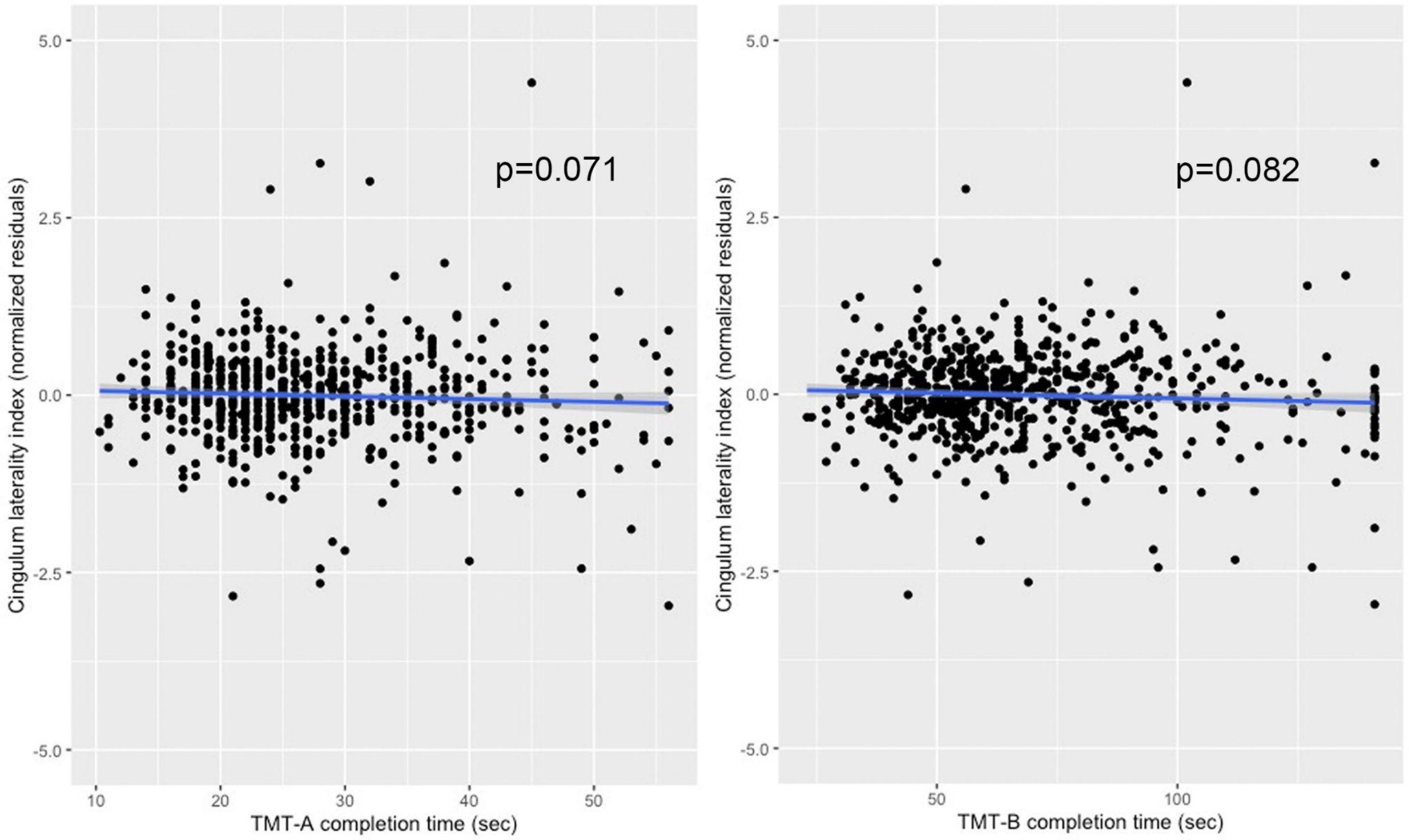

21 Meterko M, Baker E, Stolzmann KL, Hendricks AM, Cicerone KD, Lew HL. Psychometric assessment of the Neurobehavioral Symptom Inventory-22: the structure of persistent postconcussive symptoms

following deployment-related mild traumatic brain injury among veterans. *J Head Trauma Rehabil* 2012;**27**:55–62.

22 Li M, Wang M-C, Shou Y, Zhong C, Ren F, Zhang X, *et al.* Psychometric Properties and Measurement Invariance of the Brief Symptom Inventory-18 Among Chinese Insurance Employees. *Front Psychol* 2018;**9**:519.
